# Clinical exome sequencing data from patients with inborn errors of immunity: cohort level meta-analysis and the benefit of systematic reanalysis

**DOI:** 10.1101/2024.06.14.24308832

**Authors:** Emil E. Vorsteveld, Caspar I. Van der Made, Sanne P. Smeekens, Janneke H. Schuurs-Hoeijmakers, Galuh Astuti, Heleen Diepstra, Christian Gilissen, Evelien Hoenselaar, Alice Janssen, Kees van Roozendaal, Jettie Sikkema-van Engelen, Wouter Steyaert, Marjan M. Weiss, Helger G. Yntema, Tuomo Mantere, Mofareh S. AlZahrani, Koen van Aerde, Beata Derfalvi, Eissa Ali Faqeih, Stefanie S.V. Henriet, Elise van Hoof, Eman Idressi, Thomas B. Issekutz, Marjolijn C.J. Jongmans, Riikka Keski-Filppula, Ingrid Krapels, Maroeska te Loo, Catharina M. Mulders-Manders, Jaap ten Oever, Judith Potjewijd, Nora Tarig Sarhan, Marjan C. Slot, Paulien A. Terhal, Herman Thijs, Anthony Vandersteen, Els K. Vanhoutte, Frank van de Veerdonk, Gijs van Well, Mihai G. Netea, the Radboud University Medical Center multidisciplinary immune-disease board, Annet Simons, Alexander Hoischen

## Abstract

While next generation sequencing has expanded the scientific understanding of Inborn Errors of Immunity (IEI), the clinical use of exome sequencing is still emerging. We performed a cohort level meta-analysis by revisiting clinical exome data from 1,300 IEI patients using an updated *in-silico* gene panel for IEI. Variants were classified and curated through expert review. The molecular diagnostic yield after standard exome analysis was 11.8%. A systematic reanalysis resulted in the identification of variants of interest in 5.2% of undiagnosed patients, of which 75.4% were (candidate) disease-causing, increasing the molecular diagnostic yield to 15.2%. We find a high degree of actionability in IEI patients with a genetic diagnosis (76.4%). Despite the modest absolute diagnostic gain, these data support the benefit of iterative exome reanalysis in patients with IEI conveying the notion that our current understanding of genes and variants involved in IEI is by far not saturated.

## 1. Background

Inborn errors of immunity (IEI) comprise a rapidly expanding group of diseases caused by monogenic mutations that affect the function of the immune system (1,2). Patients can present a wide range of clinical symptoms including susceptibility to opportunistic infections, autoimmunity, autoinflammation, allergies, bone marrow failure and malignancy (1). Although these disorders are individually rare, the complete spectrum of IEI is substantial with more than 485 discovered IEI disease genes (1,2). The routine application of exome sequencing (ES) together with subsequent advances in sequencing chemistry, variant interpretation, and functional validation studies, have greatly accelerated IEI gene discovery (1–4). Recent discoveries of novel IEI show a trend towards the identification of autosomal dominant, *de novo* and somatic (acquired) mutations, as well as increased consideration of allelic series of existing disease genes (2). An early molecular diagnosis in any rare disease patient can prevent a long and uncertain diagnostic trajectory, guide clinical management and enable genetic counseling within the family (5).

Despite the advances in the IEI field, the yield of ES is around 30% (ranging from 15-70%) depending on cohort composition and selection, utilized *in-silico* gene panel and sequencing strategy (6,7). This gap is in part explained by the inherent technical limitations of ES, with limited detection of structural, non-coding and repeat expansion variants. In addition, variant interpretation remains challenging despite the use of diagnostic *in-silico* gene panels, that are restricted to well-established disease genes and limit the list of rare genetic variants for review. One approach to improve the diagnostic yield is reanalysis of existing genetic data, which leverages the updated knowledge base of newly described genes and genotype-phenotype correlations as well as improved variant identification, annotation and interpretation. Studies in patients with various Mendelian disorders have shown that iterative reanalysis of exome data can improve the diagnostic yield (8–14). However, the possible diagnostic gain of reanalysis of exome data in IEI patients is still under investigation (15). The rapid identification of novel disease genes and mutational mechanisms within this field specifically suggests substantial benefit of iterative reanalysis in light of new knowledge on disease genes and variants (5,16,17).

In this study, we performed a meta-analysis and investigated the diagnostic gain of systematic exome reanalysis in a large single-center cohort of patients with IEI. We systematically reanalyzed exome data at a median of 4 years after the standard analysis.

Variants were reannotated with an in-house bioinformatics pipeline that included an up-to-date *in-silico* gene panel, improved gene and variant level information, and dedicated copy number variant (CNV) analysis. Rare, non-synonymous variants in IEI genes were manually curated and classified after expert review. These data provide insights in the clinical benefit of systematic exome reanalysis for patients with inborn errors of immunity.

## 2. Methods & materials

### 2.1. Clinical patient cohort

We revisited clinical exome data from 1,300 patients from 1,272 families with a suspected IEI that were referred to the department of Human Genetics at the Radboud University Medical Center for ES between May 2013 and December 2021. Patients and their parents provided written informed consent for *in-silico* IEI gene panel analysis that is in line with the diagnostic question, as approved by the Medical Ethics Review Committee Arnhem-Nijmegen (2011/188 and 2020-7142). In addition, more than 25% of patients consented for either diagnostic exome-wide (211, 16.2%) or further academic research-based exome-wide (286, 22%) variant analysis, after counseling by a clinical geneticist. Although reanalysis of exome data could be requested *ad hoc* by the referring clinician, this was only performed in a minority of cases (N=32, 2.5%). The results of the standard analysis in a proportion of this cohort (N=253) have been previously published by our group (5), as well as reports of individual patients (18–20). Patient demographic and clinical information were available as provided by the referring clinician. Patient phenotypes were categorized according to the International Union of Immunological Societies (IUIS) for analysis and pseudonymization purposes (Table S1).

### 2.2. Standard exome sequencing

#### 2.2.1. Procedure and quality control

Sequencing was performed as previously described (5). In brief, genomic DNA samples isolated from whole blood (N=1298) or fibroblasts (N=2) were processed at the Beijing Genomics Institute (BGI) Europe (BGI Europe, Copenhagen, Denmark) or the in-house sequencing facility. All samples were enriched for coding exonic DNA using Agilent (Agilent Technologies, Santa Clara, CA, United States) or Twist (Twist Bioscience, San Francisco, CA, United States) exome kits. DNA samples at BGI were sequenced on Illumina HiSeq4000 (Illumina Sequencing, San Diego, CA, United States) or DNBseq (MGI Tech, Shenzhen, China). In-house DNA samples were sequenced on Illumina NovaSeq6000 (Illumina Sequencing). Sequencing was performed with 2×100 base pair (DNBseq) or 2×150 base pair (HiSeq4000 and NovaSeq6000) paired-end sequencing reads (Figure S1a). Sequencing quality was assessed using BamQC, which was available for 1,277 (98.2%) exomes. The mean exome coverage was 113.4x (range 65.3-373.4), the median coverage was 107.8x (Figure S1b-c).

Downstream processing was performed by an automated data analysis pipeline, including mapping of sequencing reads to the GRCh37/hg19 reference genome with the Burrows-Wheeler Aligner (BWA) algorithm and Genome Analysis Toolkit variant calling (GATK) and additional custom-made annotation (21,22). CNVs were called by copy number inference using CoNIFER while using a rolling reference pool of 500 samples (23,24).

#### 2.2.2. Systematic reanalysis

An overview of the reanalysis workflow of our study is presented in Figure 1.

**Figure 1:**
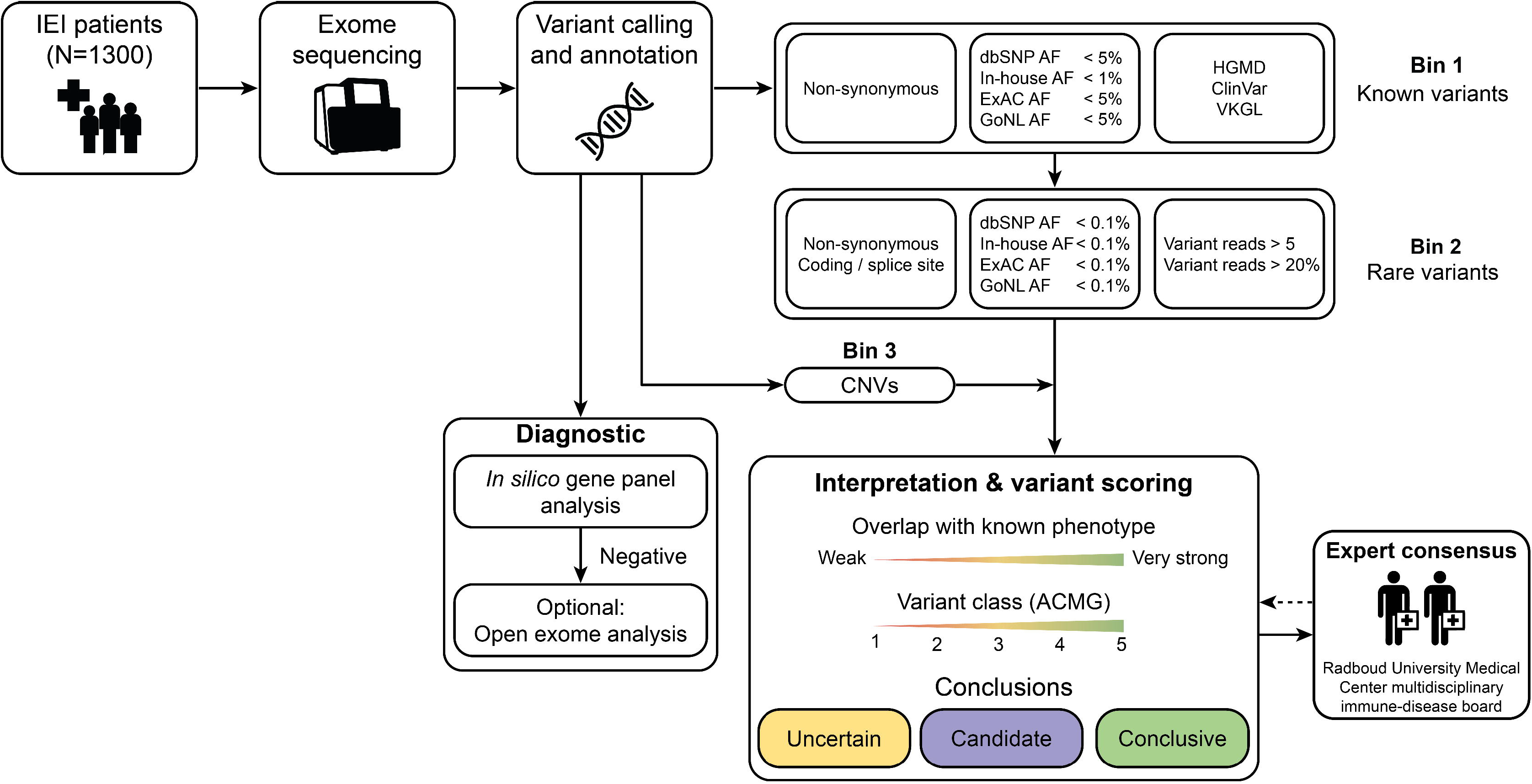
Schematic overview of the study workflow. Between 2013 and 2021, a total of 1300 patients from 1272 families with suspected inborn errors of immunity (IEI) were included in our cohort. Diagnostic exome sequencing (ES) with *in-silico* gene panel analysis was performed in all patients. Open exome analysis was undertaken in a subgroup of patients. Subsequently, the original ES data was reannotated and filtered for known pathogenic variants from variant classification databases, rare, non-synonymous coding variants and copy number variants (CNVs) that overlapped genes within the updated IEI gene panel. Filtered variants were scored based on American College of Medical Genetics (ACMG) guidelines and overlap with known gene-disease correlations and were classified into uncertain, candidate and conclusive categories following expert review.

### 2.3. Variant filtering and interpretation

For gene panel analysis, an *in-silico* filter was applied to select for variants in the IEI gene panel (25), comparable to our previous approach (5). The December 2021 IEI gene panel used at the time of the data freeze in December 2021 included 484 genes that were based on the gene lists of the International Union of Immunological Societies (1,25) with the addition of candidate genes that were proposed by a national working group but had not been published in the literature (novel candidate genes, Table S2, Figure S2) (26). Variants were further filtered using a custom Python 2.7 script using two filter strategies, referred to as bin 1 and bin 2. Bin 1 contained rare, non-synonymous variants (<5% or <1% allele frequency, respectively) in variant databases such as the Human Gene Mutation Database (HGMD) (27), ClinVar (28) and the Dutch Society of Clinical Genetic Laboratory Specialists (VKGL) Public consensus database (29). Bin 2 included non-synonymous rare coding variants (<0.1% or <0.5% allele frequency) in exons and canonical splice sites. Population databases that were used for variant filtering were dbSNP (30), ExAC (31), gnomAD (32), an in-house exome database (>50,000 exomes) and GoNL (33). All variants with variant read percentage below 20 and 5 or fewer variant reads were filtered out (Table S3). For 1,288 (99.1%) patients, CNV data called using CoNIFER was available.

Variant filtering resulted in an average of 3.44 variants in bin 1 (median 3, range 0-17, Figure S3a) and 5.42 variants in bin 2 (median 5, range 0-24, Figure S3b). These variants were independently assessed by two researchers. Variants in bin 1 were analyzed first and if no likely pathogenic or pathogenic variants were identified, bin 2 was also analyzed. Variants were prioritized by potential overlap with described phenotypes in OMIM (34), pathogenicity registered in HGMD, ClinVar or the VKGL database (where applicable) (27,28) and predicted deleteriousness pathogenicity predictors such as PhyloP and CADD (35–37).

CNV calls were filtered based on the variant call overlap with the IEI gene panel. This resulted in an average of 0.71 CNVs per patient (median 0, range 0-51, Figure S3c). Candidate CNV calls were inspected in the BAM files to assess the sequencing coverage and establish zygosity (Supplemental file 1).

### 2.4. Phenotype and variant classification

All variants were discussed in a collaborative expert meeting consisting of clinical laboratory geneticists and genome researchers to assess phenotypic overlap and variant impact. The remaining ACMG class 3-5 variants were evaluated by the Radboud University Medical Center multidisciplinary immune-disease board, which consists of diagnostic experts, clinical specialists and scientists.

The overlap of the patient phenotype with the known phenotype of a variant or gene was considered in a four-scale system, ranging from very strong (complete overlap of the clinical features with the disease spectrum in existing literature) to weak (no or minor overlap of the clinical features with the disease spectrum in the existing literature). Candidate variants were classified following the ACMG guidelines (Table S4) (29,38). Variants in patients with sufficient phenotypic overlap and a high ACMG classification (class 4/likely pathogenic and class 5/pathogenic) were considered disease-causing, hence classified as ‘conclusive’. Variants in patients with high phenotypic overlap and an ACMG VUS classification (class 3/variant of unknown significance) were considered likely disease-causing, therefore termed ‘candidate’. Variants in patients with moderate phenotypic overlap and an ACMG VUS classification were presented as ‘uncertain’ (Figure 1, Table S5). Candidate variants were counted per gene. Therefore, homozygous or compound heterozygous variants were counted as one in further statistics. More detailed clinical phenotypes from patients with candidate or conclusive causing variants after exome reanalysis was collected by the referring clinicians (Table S6).

### 2.5. Classification of reanalysis findings

We categorized the additional findings from exome reanalysis in four classes: (1) variants in genes that were added to our in-house IEI gene panel after the standard exome analysis; (2) variants in genes for which an additional disease mechanism (AR/AD/etc.) was described after the standard exome analysis, leading to reconsideration of these variants; (3) variants that are reinterpreted following new insights following literature or additional prediction software; (4) CNVs that are detected due to the addition of CNV analysis, which was not routinely performed in older cases.

### 2.6. Ancestry prediction and principal component analysis

Principal component analysis (PCA) was conducted with the smartpca program from the EIGENSOFT software package (version 7.2.1) (39). The input genotypes result from the original variant calls after applying the following filters: 1) Only variants which ‘PASS’ filter in the GATK workflow were considered; 2) Only variants with a population allele frequency in gnomAD ≥ 0.5% were considered; 3) Only variants that are located in a protein coding exon, or within 20 base pairs of a coding exon-intron boundary, were considered (based on GENCODE 31). 4) Variants were only included if they were located in regions targeted by both SureSelect AgilentV4, SureSelect AgilentV5 and TWIST exome enrichment kits. Genetic distances for all input variants were required to undertake this analysis, for which the Rutgers maps were used (40). We did not exclude any outlier in the analysis. The first two principal components are the direct output of this analysis (Figure S4, Table S7). Ancestry was determined using LASER 2.0 (41). The inferred ancestry of most patients appeared to be European (1006, 77.4%; Figure S4, Table S7).

### 2.7. TACI risk factor analysis

We gathered non-synonymous coding variants in *TNFRSF13B* in our cohort that were sufficiently covered by ES (at least 10 total reads, with >5 of the variant allele and >20% MAF). Of these, nine variants were found in patients with predominantly antibody deficiencies (PAD) and were included in the risk factor analysis. The allele frequencies in PAD patients and of the non-PAD patient within the IEI cohort were compared to a control group using the allele frequencies from gnomAD (European exomes v2.1.1) (32). Statistical differences were calculated with a Fisher’s exact test. Variants with p<0.05 after correcting for multiple testing using the Bonferroni correction were considered significant (Table S8). We further compared the allele frequency within the cohort with the allele frequency of our in-house exome database.

## 3. Results

### 3.1. Cohort characteristics

Within our cohort, the majority of patients were adult (58.6%) and female (51.9%) (Table 1). Inference of genomic ancestries predicted that 77.6% and 13.1% of patients were of the European and Middle Eastern populations, respectively (Table S7). The number of patients that were analyzed by ES increased annually from 28 in 2013 to 216 in 2021 (Figure S5). Patient classification according to the IUIS (1) indicated that the largest groups in our cohorts were autoinflammatory disorders (20,8%), diseases of immune dysregulation (17.6%) and antibody deficiencies (15.8%) (Table S1, Figure 2A).

**Table 1:**
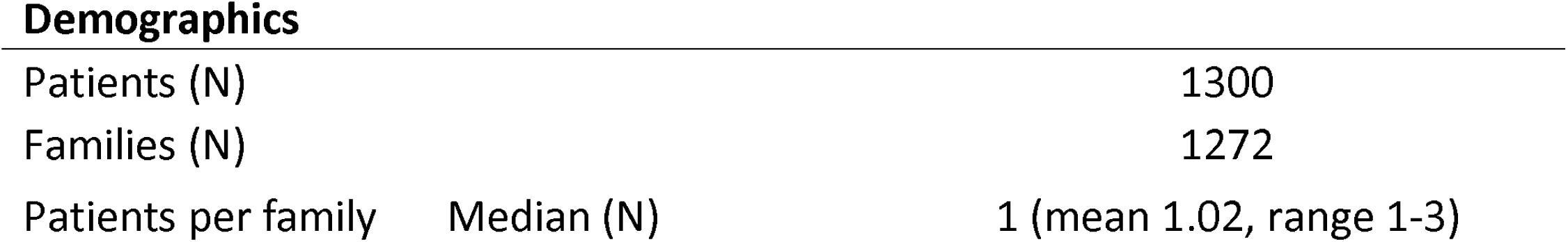
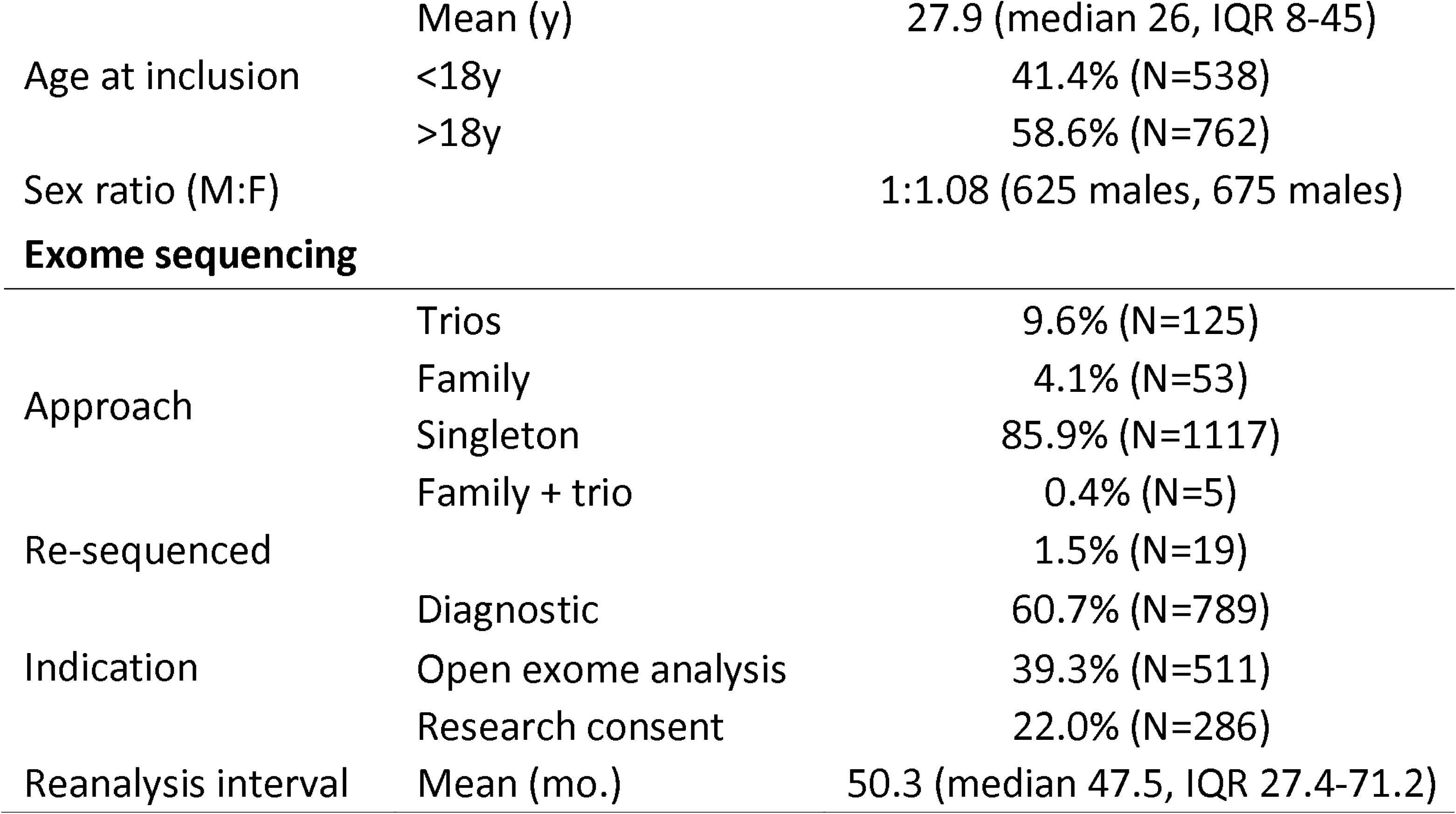
Cohort demographics and exome sequencing specifications.

**Figure 2:**
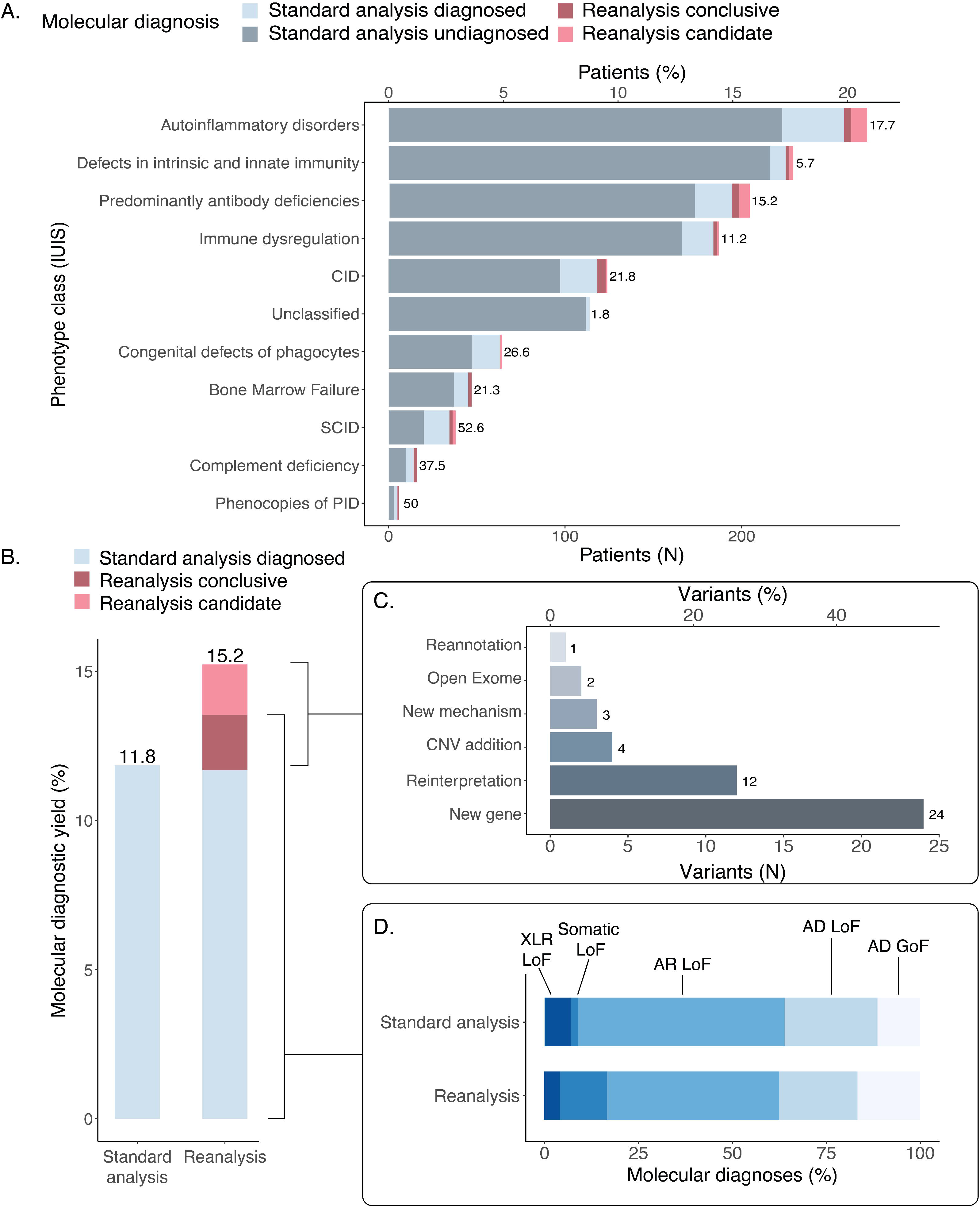
Molecular diagnostic yield after the standard exome sequencing and subsequent reanalysis in the cohort of patients with IEI. **(A)** The number and proportion of patients that received a molecular diagnosis from exome sequencing through standard analysis and reanalysis were stratified per class as defined by the International Union of Immunological Societies (IUIS). **(B)** The molecular diagnostic yield after the standard analysis and the yield of reanalysis. For two patients, an additional diagnosis was obtained through exome reanalysis. **(C)** The number and proportion of additional candidate and conclusive variants identified through reanalysis by the reason for identification. **D)** Distribution of inheritance mechanisms for the molecular diagnoses identified through standard analysis and reanalysis expressed as proportions.

### 3.2. Standard diagnostic findings

Following routine diagnostic ES, a molecular diagnosis was previously established in 154 patients from 148 families (11.8%, Figure 2B). Four patients received two molecular diagnoses (2.6%). A total of 158 diagnoses were made in these patients, of which 137 (86.7%) were established in genes included in the diagnostic IEI panel (42), and 21 (13.3%) outside the IEI panel (open exome analysis, Table S9). As expected, the diagnostic yield was higher among the patients that could be classified according to the IUIS, particularly in patients with severe phenotypes or those in categories that are supported by additional laboratory tests such as oxidative burst and complement activity in suspected phagocyte defects and complement deficiencies, respectively (Table S1). Within the full cohort, 114 patients could not be classified based on their phenotype. Only two of these patients received a molecular diagnosis (1.8%). Additionally, the diagnostic yield was higher for pediatric patients compared to adult patients (18.2% and 7.3%, Table S10). Of the previous molecular diagnoses, most followed autosomal recessive inheritance (n=87, 55.1%), followed by autosomal dominant (n=57, 36.1%) and X-linked recessive (n=11, 7.0%) inheritance, while only 3 diagnoses followed from somatic mutations (1.8%) (Figure 2D). The most frequent molecular diagnoses included mutations in *MEFV* (n=8), *NFKB1* (n=5), *DNMT3B* (n=5) and *ADA2* (n=5) (Table S9).

### 3.3. Diagnostic yield of exome reanalysis

Exome reanalysis was performed at a median of 47.5 months following the standard analysis (Table 1, Figure S6). Each variant was scored based on gene and variant level metrics and genotype-phenotype correlation and subsequently classified following expert review (Figure 1). Targeted reanalysis of variants in the latest version of our in-house IEI gene panel in our cohort of 1,300 patients resulted in the identification of 61 relevant variants in 60 out of 1,146 undiagnosed patients (5.2%). This reanalysis led to a definitive molecular diagnosis in 24 (2.1%) patients, of which two patients received an additional diagnosis. We further identified candidate variants in 22 patients (1.9%), which were VUS that were found in patients exhibiting phenotypic features that had substantial overlap with the known diseases associated with mutations in these genes. Together this increased the (candidate) molecular diagnostic yield from 11.8% of the initial analysis to 15.2% (Table 2-3, Figure 2B, Table S11). Subsequently, we identified 15 patients with a VUS in a gene that could be related to the patient’s phenotype (1.3%) and were therefore classified to be of uncertain clinical relevance (Table S5). Of all these variants, 55 were single nucleotide variants (SNV) (90.2%) and 6 were CNVs (9.8%). The variants that were identified during reanalysis were subsequently categorized into six classes based on the reason for identification: 1) variants in a new disease gene, 2) known variants of which the effect was re-interpreted based on new insights from mutation databases, literature, and prediction software, 3) variants detected after dedicated CNV analysis, 4) variants associated with a novel mutational mechanism, 5) previous open exome analysis and 6) variant reannotation (Figure 2C, Table 1-2, Table S12-13).

**Table 2:**
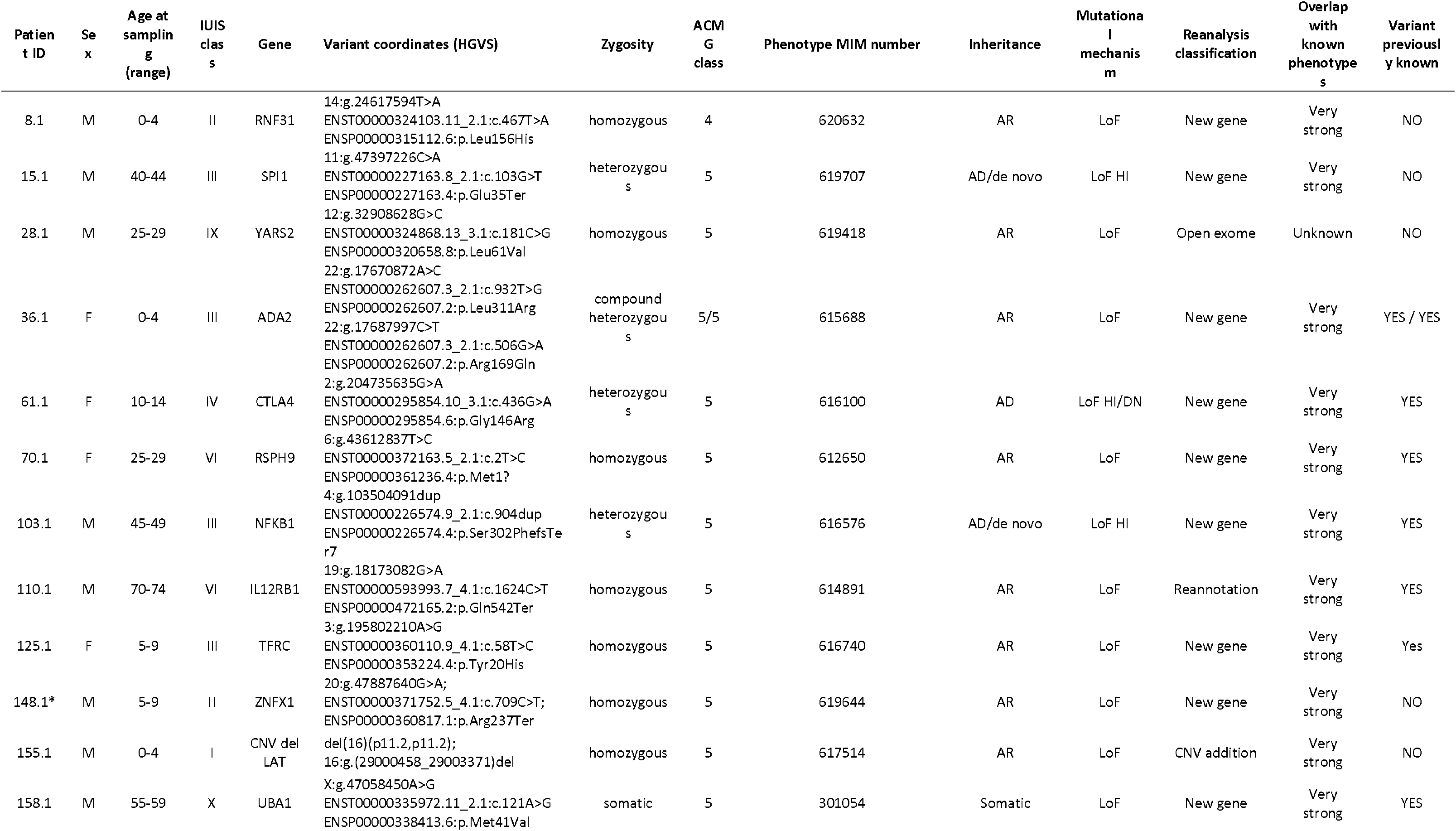
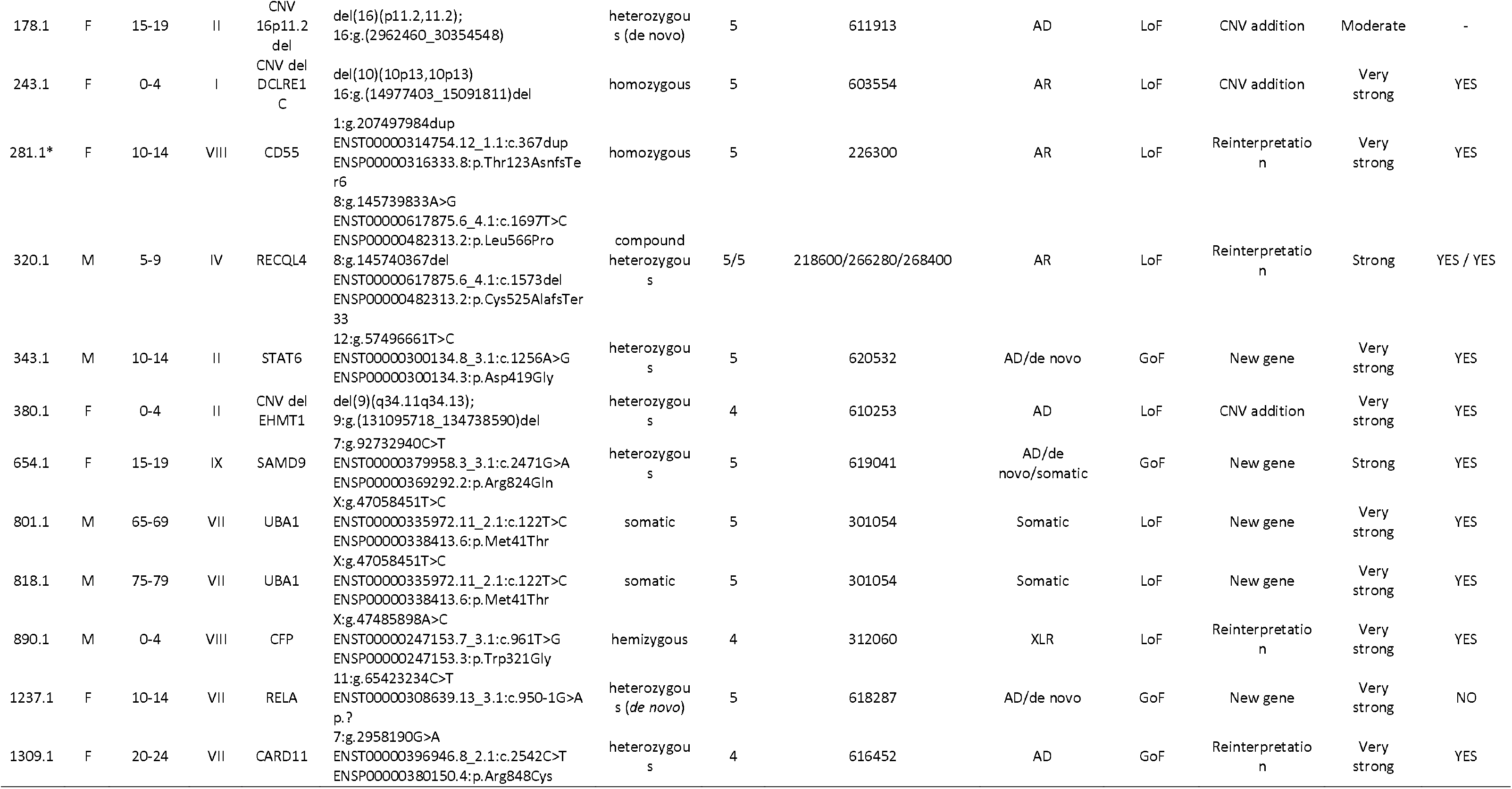
Patients diagnosed through exome re-analysis. Variants identified through reanalysis that led to a conclusive molecular diagnosis. ^*^Patient received an additional diagnosis (Table S9).

#### 3.3.1. Novel IEI genes

In 28 of the 60 probands (46.7%) for which additional variants were identified, these were found in new IEI genes for which the disease associations were characterized after the standard analysis. Of these, 24 probands carried candidate or conclusive variants that led to a (likely) diagnosis (Figure 2C, Table 2-3, Table S11). Most variants were considered in genes that followed AD (*IRF2BP2* (n=2), *CTLA4, SPI1, NFKB1, OAS1, SOCS1, STAT6, SAMD9, RELA* (*de novo*)) and AR (*ADA2, RNF31, RSPH9, TFRC, CHUK, ZNFX1, SLC39A7, TET2*) inheritance patterns, but also XLR (*SASH3*) and acquired somatic (*UBA1* (n=3)). These variants were documented in clinical databases as (likely) pathogenic (*NFKB1, SAMD9, SAMD9L, ADA2, RSPH9, UBA1, TFRC*), exerted genetic loss-of-function (*IRF2BP2, SPI1, NFKB1, RSPH9, ZNFX1, TFRC, RELA*), approximated amino acid positions of published mutations with similar predicted effects (*SLC39A7*), and/or showed convincing phenotypic overlap consistent with the literature (*IRF2BP2, OAS1, SOCS1, CHUK, SASH3, TET2*). In addition, the *STAT6* variant was found to confer a gain-of-function effect and was extensively characterized as part of a larger effort (43). The patients with *RELA* and *UBA1* variants were also published previously (19,44).

#### 3.3.2. Variant re-interpretation

A total of 19 patients of the 60 patients (31.7%) for which we identified additional variants in were found to be present in genes that were previously part of the standard *in-silico* gene panel, but were reclassified based on recent insights, of which 12 were candidate or conclusive variants. In 5 probands, previous VUS were classified upward to (likely) pathogenic based on documentation in clinical databases (Table 2-3, Table S11). Based on internal expert review and additional case descriptions in the literature, another 6 probands had variants that were re-interpretated as candidate variants (Table 3). For example, proband 1317.1 carried a heterozygous missense variant in *NFKB2* (p.(Arg193His)) that was not present in any population database and had high scores for deleteriousness (i.e. CADD-PHRED 26.9), matching his CVID phenotype that was complicated by granulomatous lymphocytic interstitial lung disease (GLILD) (45).

**Table 3:**
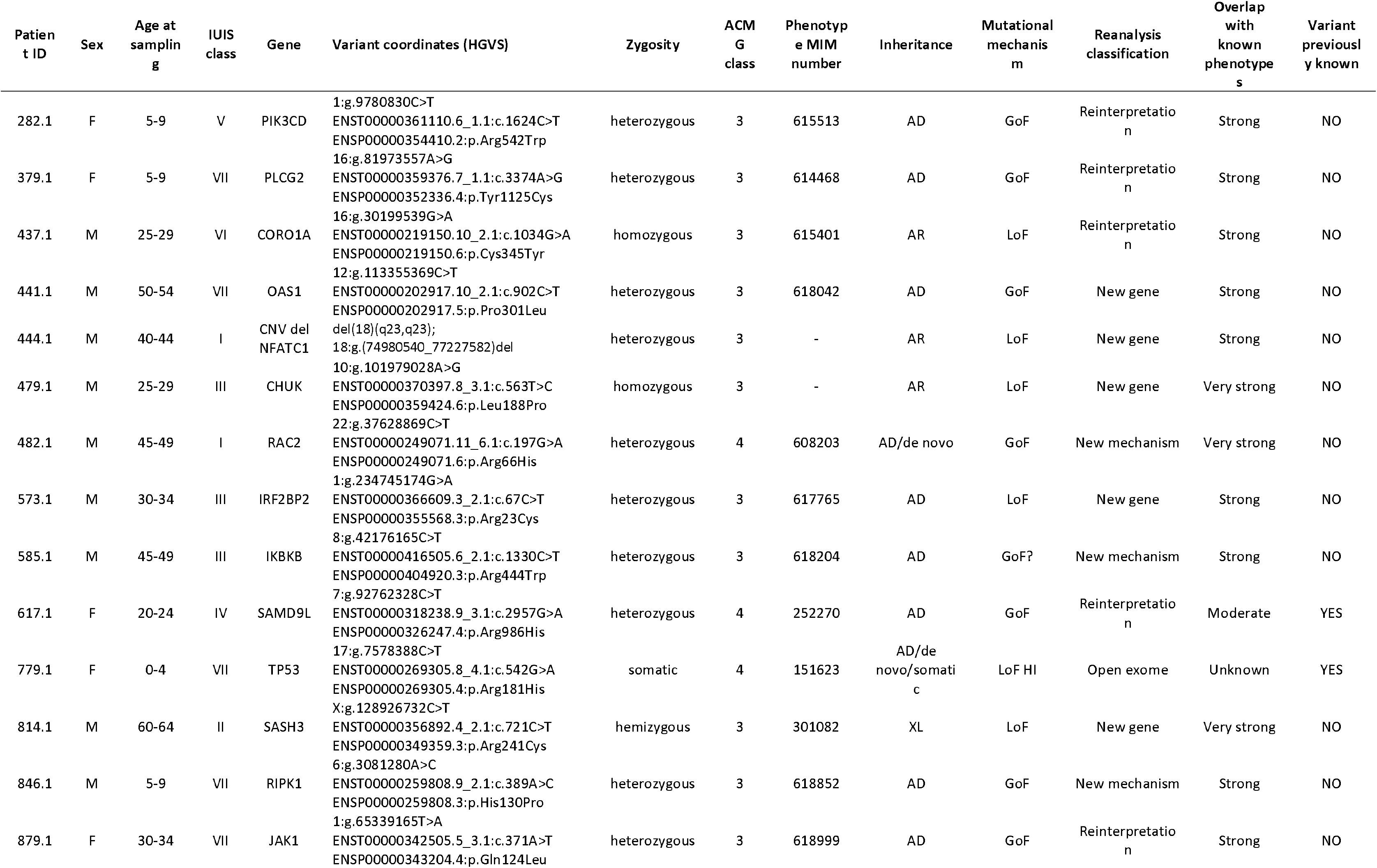
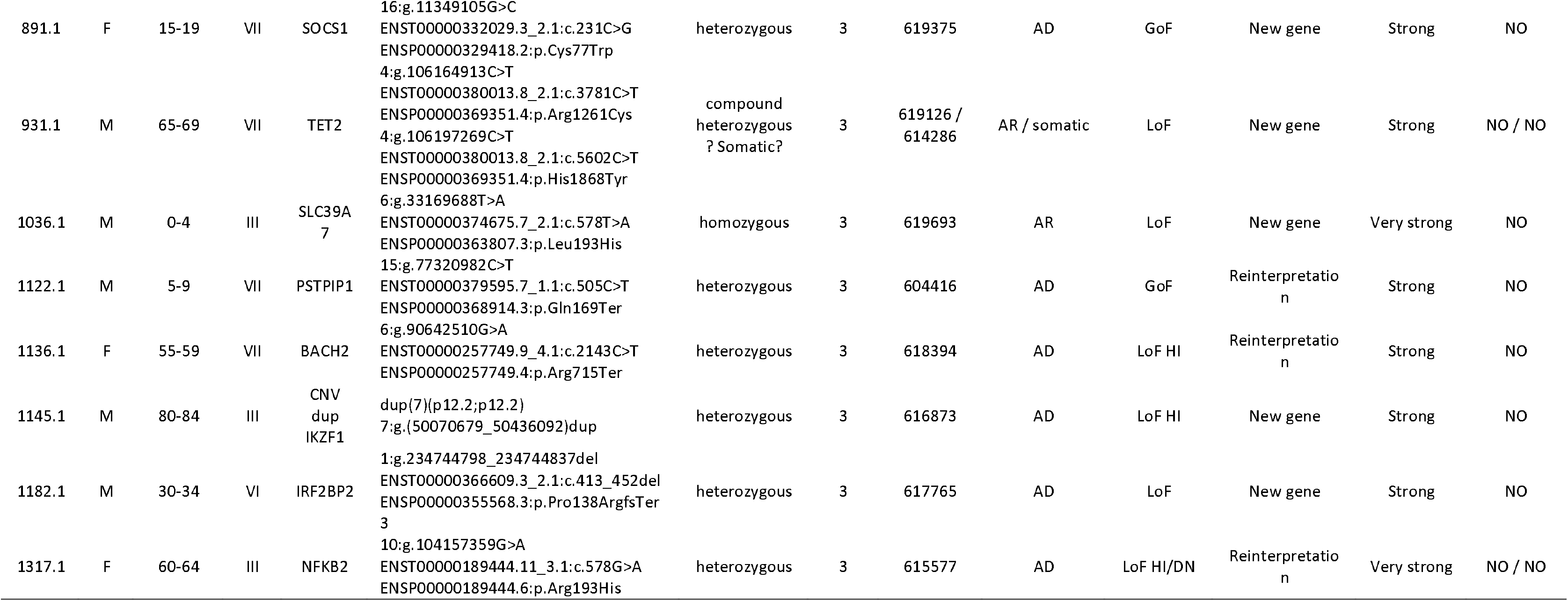
Cases with novel candidate variants after reanalysis. Variants identified through exome reanalysis that are classified as VUS in patients where the phenotype overlaps significantly with those described in literature.

#### 3.3.3. Additional CNVs

Systematic assessment of CNVs returned six CNVs that overlapped IEI genes, of which four were novel and considered conclusive and two were previously called but overlapped our updated gene panel (*IKZF1, NFATC1;* Table 3, Supplemental File 1). Proband 155.1 carried a homozygous 16p11.2 deletion that was confirmed by XON array and was shown to overlap the *LAT* locus. This led to the conclusion that LAT deficiency was the explanation of the patient’s T-B+ SCID phenotype. A de novo 16p11.2 deletion was detected in proband 178.1, who had documented symptoms of immunodeficiency, neutropenia and a speech disorder. A homozygous 10p13 deletion that includes the *DCLRE1C* locus was detected in proband 243.1, supporting the diagnosis of Artemis deficiency. Finally, proband 380.1 had a 9q34 microdeletion that has been described in patients with Kleefstra syndrome through haploinsufficiency of *EHMT1*, a neurodevelopmental disorder associated with recurrent infections that resembled the probands phenotype (46).

#### 3.3.4. Novel mutational mechanisms

Three probands (5.0%) had candidate or conclusive variants in known IEI genes with recent discoveries of novel mutational mechanisms. Patient 482.1 with CID harbored a missense variant in RAC2 (p.(Arg66His)) that was close to a known gain-of-function mutation (p.(Glu62Lys)) identified in six patients with the same phenotype (47,48). Moreover, the heterozygous *IKBKB* variant (p.(Arg444Trp)) in patient 845.1 has also been reported in association with the same phenotype of CVID (49), while AR *IKBKB* mutations were initially described to underlie SCID (50,51), but recently also as AD gain-of-function in CID (52). Lastly, we observed a heterozygous missense *RIPK1* variant (p.(His130Pro)) in patient 846.1 with autoinflammatory disease. This variant was not located near the caspase 8 cleavage site that is a hotspot for heterozygous mutations in cleavage-resistant RIPK1-induced autoinflammatory disorders (CRIA). The possible mutational mechanism is therefore unclear, but it could be hypothesized that its location in the kinase domain near the necrostatin binding site could impact the activation loop and lead to an increase of RIPK1-dependent NF-kb activation, apoptosis and necroptosis (53).

#### 3.3.5. Actionability

We evaluated potential clinical actionability by cross-referencing the molecular diagnoses with curated gene-disease lists from the ClinGen database (54) and a comprehensive previous review (55), supplemented by recent literature (Figure S7, Table S13). In a subset of the current cohort, we have previously reported targeted therapeutic options in 76.4% of patients with a molecular diagnosis based on expert opinion and literature review (5). Analysis of the current cohort indicated that 123 out of 154 patients had a molecular diagnosis (79.9%) that was associated with ≥ 1 clinical management options, including supportive (n=118, 76.6%), preventative (n=94, 61.0%) or targeted (n=62, 40.2%) therapy, hematopoietic stem cell therapy or other organ transplantation (n=59, 38.3%), or gene therapy (n=8, 5.3%). Similarly, 20 out of 24 (83.3%) probands had a novel molecular diagnosis after reanalysis that was considered actionable, suggesting options for supportive (n=19, 79.2%) or preventative (n=17, 70.8%) therapy, hematopoietic stem cell therapy (n=11, 45.8%), targeted therapy (n=8, 33.3%), or gene therapy (n=1, 4.2%).

### 3.4. Risk factors for antibody deficiency

Leveraging the size of the current cohort, we studied the prevalence of variants in the gene encoding for TACI (*TNFRSF13B*) that are considered risk factors for antibody deficiency (Figure 3, Table S8). Specifically, we collected all non-synonymous coding variants and compared the allele frequency with those reported in the presumably healthy population in gnomAD and our in-house database (32). We identified enrichment of six *TNFRSF13B* variants, of which five variants were found at higher allele frequencies in our cohort, and one variant (p.(Pro251Leu)) was found at a higher allele frequency in gnomAD (Figure 3A). Four variants were present at a higher AF in patients with PAD compared to gnomAD (p.(Arg20Cys), OR=82.1 CI 9.20-346.93; p.(Cys104Arg), OR=17.7; p.(Leu171Arg), OR=85.4; p.(Ala181Glu), OR=7.4, Figure 3B). In the full cohort, we found 66 patients (5.1%) that carried a previously described risk variant in *TNFRSF13B* (p.(Cys104Arg), p.(Leu171Arg) and p.(Ala181Glu)), of which 38 patients carried the most common p.(Cys104Arg) variant (3 homozygotes, 35 heterozygotes; 2.9%). Conforming to ample literature describing an enrichment of these three variants in patients with common variable immunodeficiency (CVID), we found a higher carrier proportion among patients with PAD compared to the rest of the cohort (31/204, 15.2%), including 19 carriers of p.(Cys104Arg) (9.3%, 17 heterozygotes, 2 homozygotes) (56–58). In addition, the p.(Arg20Cys) variant that was identified in 3 individuals (two with PAD/CVID, one with autoinflammation) has not been characterized as a risk factor for PAD but has been put forward as a candidate variant (59– 61). Of the 34 PAD patients that carried a risk variant, antibody deficiency was confirmed in 29 patients (93.5%) based on additional clinical information. None of the carriers of these four variants among PAD patients had received an alternative, previous molecular diagnosis.

**Figure 3:**
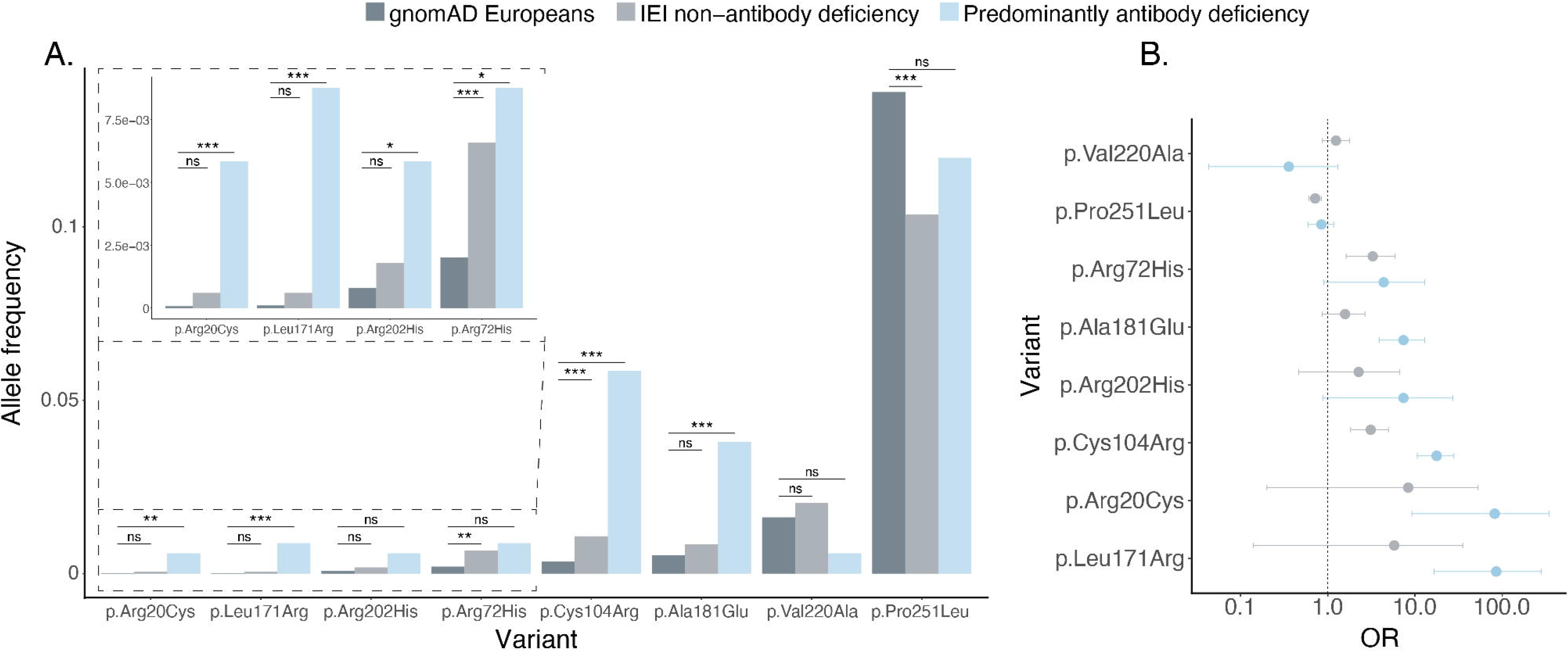
Enrichment of TNFRSF13B risk factors for antibody deficiency in our cohort. **(A)** Comparison of the allele frequencies of non-synonymous coding *TNFRSF13B* variants in patients with and without a primary antibody deficiency versus the European GnomAD population database. Statistically significant results are indicated with asterisks (*: p <0.05, **: p <0.01, ***: p <0.001) and were calculated using a Fisher’s exact test. **(B)** Visualization of calculated odds ratios for these variant allele frequencies in the different populations with 95% confidence intervals.

Of the four alleles that were enriched in PAD patients, the p.(Cys104Arg) variant was also significantly enriched in the non-PAD classified patients within our cohort when compared to the GnomAD and in-house database AF, although at a significantly lower AF compared to the PAD patient group (Table S8). Interestingly, review of the clinical phenotype of the non-PAD patients showed that 7/19 carriers did have a documented antibody deficiency, 6/19 carriers had normal immunoglobulin levels and 6/19 carriers were not tested. These results indicate a significant correlation between a TACI risk factor and antibody deficiency in IEI patients and propose the p.(Arg20Cys) variant for further investigation as a risk factor for PAD/CVID.

## 4. Discussion

Exome sequencing is a valuable and cost-effective first-tier genetic test for the molecular diagnosis of IEI (5). The rapidly increasing knowledge on new IEI disease genes and mutational mechanisms paired with improved bioinformatic analysis pipelines suggests potential benefit of iterative exome reanalysis, but this has so far not been comprehensively investigated. In this study, we report on the standard diagnostic yield and the gain of systematic exome reanalysis in our single-center cohort of 1,300 IEI patients after a median reanalysis time interval of 4 years. We generated updated gene- and variant level information for SNVs and CNVs that comprised an expanded *in-silico* IEI gene panel and multiple strict filtering steps were used to limit the analysis workload. Manual curation led to the identification of additional variants in 5.2% of undiagnosed patients (N=60), of which 75.4% were considered candidate or conclusive variants after expert review. Most of the identified variants were found in new disease genes, re-interpreted based on literature and variant databases, associated with novel mutational mechanisms, or identified by dedicated CNV analysis. Our reanalysis increased the (candidate) molecular diagnostic yield in the complete cohort by 28.6%, from 11.8% to 15.2%. We further highlight the actionability in diagnosed IEI patients, with 83.3% of patients with a diagnosis from exome reanalysis that was considered actionable. Despite the modest diagnostic gain achieved through exome reanalysis, these data support the application of iterative reanalysis to aid in the genetic diagnosis of patients with IEI. This study demonstrates that both gene- and variant level knowledge for IEI is still steadily increasing, possibly more than for other rare genetic disorders with a higher molecular diagnostic yield (62). Additionally, our study indicates the importance of a dialogue between experts in the diagnosis of patients with IEI, and the benefit of variant evaluation by specialists from various disciplines in the evaluation of variants identified through exome reanalysis.

### 4.1. Results of previous exome reanalysis efforts

Reanalysis of genomic data can be an effective method to solve undiagnosed patients. Various studies have described reanalysis efforts in patients with rare Mendelian diseases, with notable differences in cohort size, patient characteristics, reanalysis interval, and standard sequencing and reanalysis approaches. We have summarized the most important characteristics of these studies after a non-exhaustive literature review (Table S14). We included 38 studies that describe the reanalysis of ES and/or GS sequencing data from cohorts of rare disease patients. All studies were published between 2017 and 2023 and included varied patient cohorts with between 12 and 4,411 individuals (mean 520, median 166). Reanalysis approaches most often included *in-silico* gene panel updates and variant reannotation, but also additional phenotyping, bioinformatics approaches and the application of additional genome-wide and targeted sequencing approaches. In general, larger cohort studies reported additional diagnostic yields ranging between 2-20%. The benefit was greater in studies with patients with neurodevelopmental disease (NDD), more trio- or family-based sequencing data, and a longer reanalysis interval.

Only two previous studies have endeavored reanalysis in an IEI patient population. A small study with 94 IEI patients reported no additional molecular diagnoses after performing a gene panel update almost 23 months after the standard ES (15). Most recently, a large study from the National Institute of Allergy and Infectious Diseases (NIAID) with a cohort of 1,000 probands from the 1,505 kindreds found that reanalysis led to 3.3% novel molecular diagnoses at a median of almost two years (15.1-34.0 months) after the first analysis that had provided a molecular diagnosis in 327 families (32.7%) (63). The standard diagnostic yield of this study was substantially higher than in our cohort, which could be attributable to differences in the studied population, analysis approach and variant interpretation. Although our cohort was selected based on the availability of exome data, this cohort contained a quarter of patients that already had a diagnosis before referral. The authors further report that almost three-quarters of molecular diagnoses reflected specific patient populations of interest to the NIAID. In addition, their analysis approach was not limited to an *in-silico* gene panel, resulting in 8.3% patients with a non-immune diagnosis, and additional solved cases after microarray analysis. Lastly, risk factors such as *TNFRSF13B* were also considered as molecular diagnoses, similar to other studies that employ NGS for the diagnosis of IEI, and secondary findings were also counted (63–65). The reanalysis in this study was comprised of an update of new disease-gene associations, variant reclassification, segregation analysis and improved CNV detection with microarray analysis (CMA). Similar to our findings, most new diagnoses were accounted for by updated disease-gene associations and variant reinterpretation, due to upward classification and additional clinical data, and improved CNV detection. Interestingly, only 13 of 22 new diagnoses (59.1%, 1.9% of the total of undiagnosed patients) were represented in the IUIS list. In our cohort, we only identified 3 of 44 candidate or conclusive variants in genes outside of the IUIS gene list, since we could evaluate exome-wide variants in only a small proportion of patients. These studies suggest that targeted reanalysis with a median interval of 2-4 years could improve the diagnostic gain by ∼3% in patients with diverse IEIs with a potential benefit of exome-wide evaluation, but at the cost of more variants of unknown significance and secondary findings.

### 4.2. The potential for studying risk factors and modifiers

TACI deficiency is the most common genetic defect in up to 10% of CVID patients, caused by variants in *TNFRSF13B* that follow autosomal dominant or recessive inheritance. (1,57,66). Moreover, it has been shown that the presence of a TACI risk factor correlates with lymphoproliferation and autoimmune manifestations (62). These TACI risk variants are present at relatively high allele frequencies in the general population (Table S8), highlighting the considerable variability in clinical expression and penetrance of these variants (67,68). Functional studies have shown that these *TNFRSF13B* variants interfere with signaling through the TACI receptor through impaired ligand binding, receptor oligomerization and downstream MYD88 signaling, thereby disrupting its function in B cell homeostasis, isotype switching and antibody responses (69). Within our cohort, we replicated known enrichment of *TNFRSF13B* variant alleles in patients with PAD. We also found an enrichment of the p.(Cys104Arg) variant in non-PAD patients, which included patients that did have antibody deficiency as a secondary diagnosis. Furthermore, we identified enrichment of the p.(Arg20Cys) (rs200013015) variant in PAD patients (OR=82.12 CI 9.20-346.93; p=0.0028), which has previously been classified as VUS in a patient without an immune-related phenotype (59), after which it was also identified in patients with antibody deficiency (60,61). Despite a significant statistical enrichment as demonstrated here, this variant is predicted to have a benign consequence on protein function. Therefore, further functional studies are needed to investigate its effect on B cell function (Table S8). Patients with PAD and a TACI risk factor had no alternative molecular diagnoses, supporting the strong genotype-phenotype correlation and relevance of reporting these (relatively) common variants in diagnostics. The expanding size of rare disease cohorts through routine genetic testing and international data sharing holds promise for the investigation of non-monogenic inheritance patterns, including the contribution of risk factors and modifiers. As demonstrated here by calculating odds ratios per variants in *TNFRSF13B*.

### 4.3. The future of ES reanalysis in the IEI field

Several lessons can be learned from the multitude of reanalysis approaches that have been undertaken to demonstrate its diagnostic benefit.

First, reanalysis should be periodically and systematically performed or can be initiated *ad hoc* by the patient or treating physician. The reanalysis interval should hold a balance between the improvement of variant annotation, increase in disease-gene knowledge and expected molecular yield versus cost of the required time investment. Although the diagnostic gain of our study after almost four years of follow-up was modest, the rapidly growing number of novel IEI genes and novel mutational mechanisms of known genes would advocate for relatively frequent reanalysis (1,2). Also in our cohort, a significant proportion of candidate variants (N=3, 6.5%) were observed in known genes with novel inheritance patterns, in particular AD disease caused by gain-of-function (Figure 2D, Tables 2-3, Table S12). We further observed that variants identified through reanalysis were more often autosomal dominant gain-of function variants (16.7% compared to 11.7%), whereas variants found after the initial analysis were more likely to be autosomal recessive loss of function variants (55.1% compared to 45.8%, Figure 2D). This trend results from the increased proportion of AD (GoF) disease gene mechanisms among new IEI gene discoveries. Most studies describing exome reanalysis have suggested annual or biannual intervals, in part depending on the workload and level of automation (9,11,13,70–72). Fully automated reanalysis strategies have been described, for example by using software- or artificial intelligence-based variant classification or interpretation (73,74), or an application programming interface (API) that integrates genome and phenome data (75). Future advances in bioinformatics infrastructure could facilitate frequent automated reannotation of available genomic data and integration with systematic phenotyping data, thereby prioritizing variants for expert review.

Second, the reanalysis strategy can implement additional sequencing, variant filtering or ancillary technologies to enhance the molecular diagnostic yield. ES-based resequencing with superior advancements in sequencing chemistry compared to outdated exome data can significantly upgrade data quality and is recommended by medical genetics guidelines (12,14,76,77). Moreover, the application of trio-ES has been shown to improve variant prioritization and interpretation, allowing for segregation and variant phasing in the affected proband and potentially affected family members (12,20,72,78,79). Variant filtering can also be adjusted to improve detection of other variant types such as somatic mutations. Other ancillary technologies such as genome sequencing (GS), long read sequencing (80) and optical mapping could be employed to uncover genetic variation that was missed in ES data, including CNVs, large structural variants and non-coding variants (81–84). The diagnostic yield of short-read GS has been shown to be limited, although with declining prices it may be foreseeable that GS rather than ES will become the first-tier approach, even if the primary focus remains on coding variants or exome-from-genome analyses (79,85,86). The future move towards long-read GS in clinical genetics could constitute a one-test solution that informs on the most clinically relevant genetic variation (87). Lastly, omics-based technologies such as (single cell) transcriptomics, proteomics, metabolomics and methylation profiling are emerging and an integration of iterative reanalysis with these approaches may be opportune specifically for the study of IEI patients, as affected cells are usually more easily accessible. Antibody-based cytometry techniques could additionally offer complementary approaches to discover the underlying mutation for specific IEI patients (83).

Third, data sharing and analysis at the national (26,88) and international (89) level can improve the power to uncover novel molecular diagnoses in large IEI disease cohorts. International consortia such as the pan-European Solve-RD project are in the process of conducting large-scale systematic reanalysis efforts of existing exome and genome datasets across multiple rare disease fields that will create more standardized and scalable frameworks (90). Large-scale analyses could also enable the potential for investigating non-

Mendelian inheritance, such as digenic or oligogenic inheritance and incomplete inheritance in rare diseases such as IEI (91–93). Moreover, data sharing through platforms such as GeneMatcher (94) and Matchmaker Exchange (95) could assist in the identification of novel genotype-phenotype correlations.

### 4.4. Limitations

This study has several limitations. First, the findings within our cohort might not be extrapolated to other IEI populations (63,65,96). The molecular diagnostic rate in our cohort is limited, mainly owing to patient characteristics, including a predominantly adult population with late disease onset, high proportion of disease categories with a lower chance of monogenic findings, and the inclusion of less severe immune phenotypes where ES is used to rule out an IEI. The distribution of patient phenotypes differs from those in the large patient registry for IEI from the European Society for Immunodeficiencies, which includes a higher percentage of patients with PAD and lower percentages of patients with immune dysregulation, defects in intrinsic immunity and autoinflammatory disease (97). Nevertheless, our cohort reflects clinical practice with an unselected, cross-sectional cohort and a wide array of disease manifestations. Therefore, the reported standard molecular diagnostic yield and benefit of systematic reanalysis could be considered as representative for centers with mixed IEI populations. Second, to provide a comprehensive overview of our study population, patients were classified according to the IUIS, which is intended for established molecular diagnoses and could introduce bias in case of limited or inaccurate phenotypic information. Systematic phenotyping could improve patient classification, for instance by using the Human Phenotype Ontology, and could facilitate phenotype-driven exome reanalysis (72,75,98). This would not affect the risk factor analysis, since patients with PAD could be reliably classified based on the available phenotypic information. Lastly, the retrospective design of our study might introduce bias in the reinterpretation of exome data. A prospective study with an iterative semi-automated analysis and systematic clinical phenotyping might offer a more optimal and unbiased evaluation of the diagnostic value of reanalysis and the direct benefits for patient care.

### 4.5. Conclusion

Our results support the diagnostic benefit of ES reanalysis in a cohort of 1,300 suspected IEI patients after a four-year follow-up interval. We reannotated ES data with updated gene- and variant level information for SNVs and CNVs and prioritized variants in the latest version of the IEI gene panel based on expert review. We identified additional variants in 5.2% of undiagnosed patients, increasing the (candidate) molecular diagnostic yield from 11.8% to 15.2%. Most novel variants were found due to new disease-gene associations, variant re-interpretation or systematic CNV analysis. Furthermore, we found enrichment of TACI risk factor variant alleles in patients with antibody deficiency. Lastly, we propose possibilities to maximize the clinical benefit of future reanalysis studies.

## Data Availability

The datasets supporting the conclusions of this article are included within the article and supplementary files. All raw data was generated within the diagnostic procedure and analysis. This does not allow public data sharing as these data can be identifiable information, for which the patients or their families did not provide consent.
All identifiable information has been redacted. Patient IDs used within the study are only known within the research group.

## List of abbreviations

AD: Autosomal dominant
AR: Autosomal recessive
CNV: Copy⍰number variant
ES: Exome sequencing
GoF: gain-of-function
GS: Genome sequencing
IEI: Inborn errors of immunity
LoF: loss-of-function
NGS: Next generation sequencing
PCA: Principal component analysis
SNV: Single nucleotide variant
VUS: Variant of unknown significance
XLR: X-linked recessive

## Ethics approval and consent to participate

Patients/families were counseled and provided informed consent in the realm of the diagnostic procedure.

## Consent for publication

Not applicable

## Availability of data and materials

The datasets supporting the conclusions of this article are included within the article and supplementary files. All raw data was generated within the diagnostic procedure and analysis. This does not allow public data sharing as these data can be identifiable information, for which the patients or their families did not provide consent.

## Authors’ contributions

Conceptualization: EEV, CIM, AS, AH; data curation: EEV, CIM, GA; formal analysis: EEV, CIM, WS; investigation: EEV, CIM, SM, AS, HD, EH, AJ, KvR, JSvE, TM, MMW, AH; resources, including patient recruitment and referral: JSH, GA, WS, MSA, KA, BD, EAF, SSVH, EH, EI, TBI, MCJJ, RK, IK, ML, CMM, JO, JP, NTS, MCS, PAT, HT, AV, EKV, FV, GW; software: EEV; supervision: MGN, AS, HY, CG, AH; visualization: EEV, CIM; writing—original draft: EEV, CIM; writing—review and editing: All authors. All authors read and approved the final manuscript.

### Acknowledgements

We thank all patients and their families for their contribution to this study. We would also like to thank all colleagues from the diagnostic division of the Human Genetics Department, from the Radboud Genomics Technology Center and the Radboud University Medical Center multidisciplinary immune-disease board for their support.

## Table and figure legends

### Additional Figures

**Figure S1:**
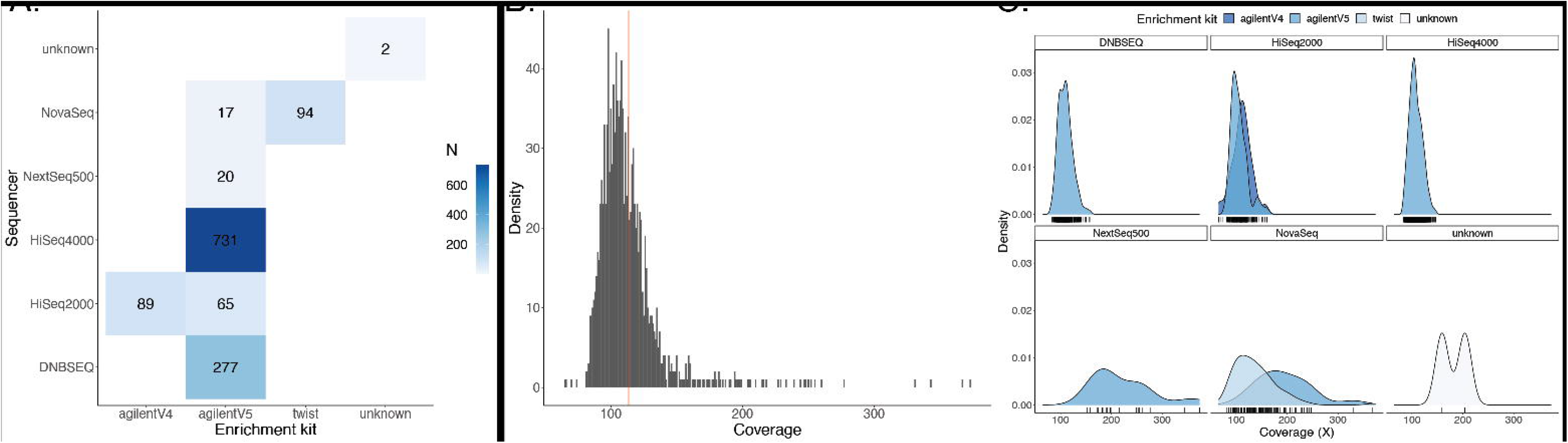
Technical details of the exome sequencing procedure in our cohort. **A** Combination of exome enrichment kit and sequencer. **B** Sequencing coverage distribution. **C** Coverage per exome kit-sequencer combination. The median number of variants per patient is indicated with a red vertical line.

**Figure S2:**
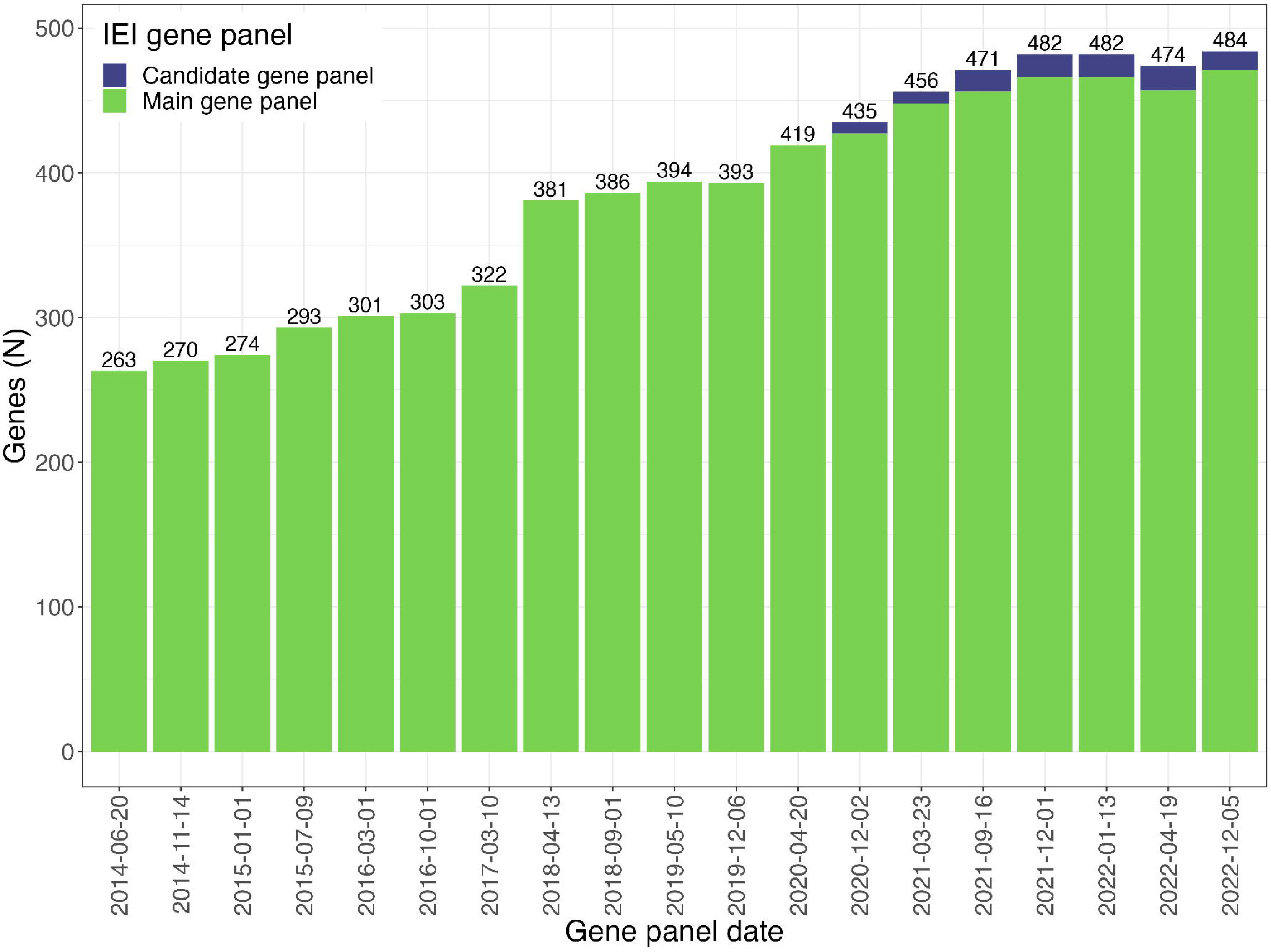
Expansion of the in-house inborn errors of immunity gene panel. The number of genes in the main panel and all candidate genes (novel candidate disease genes) in green and blue, respectively.

**Figure S3:**
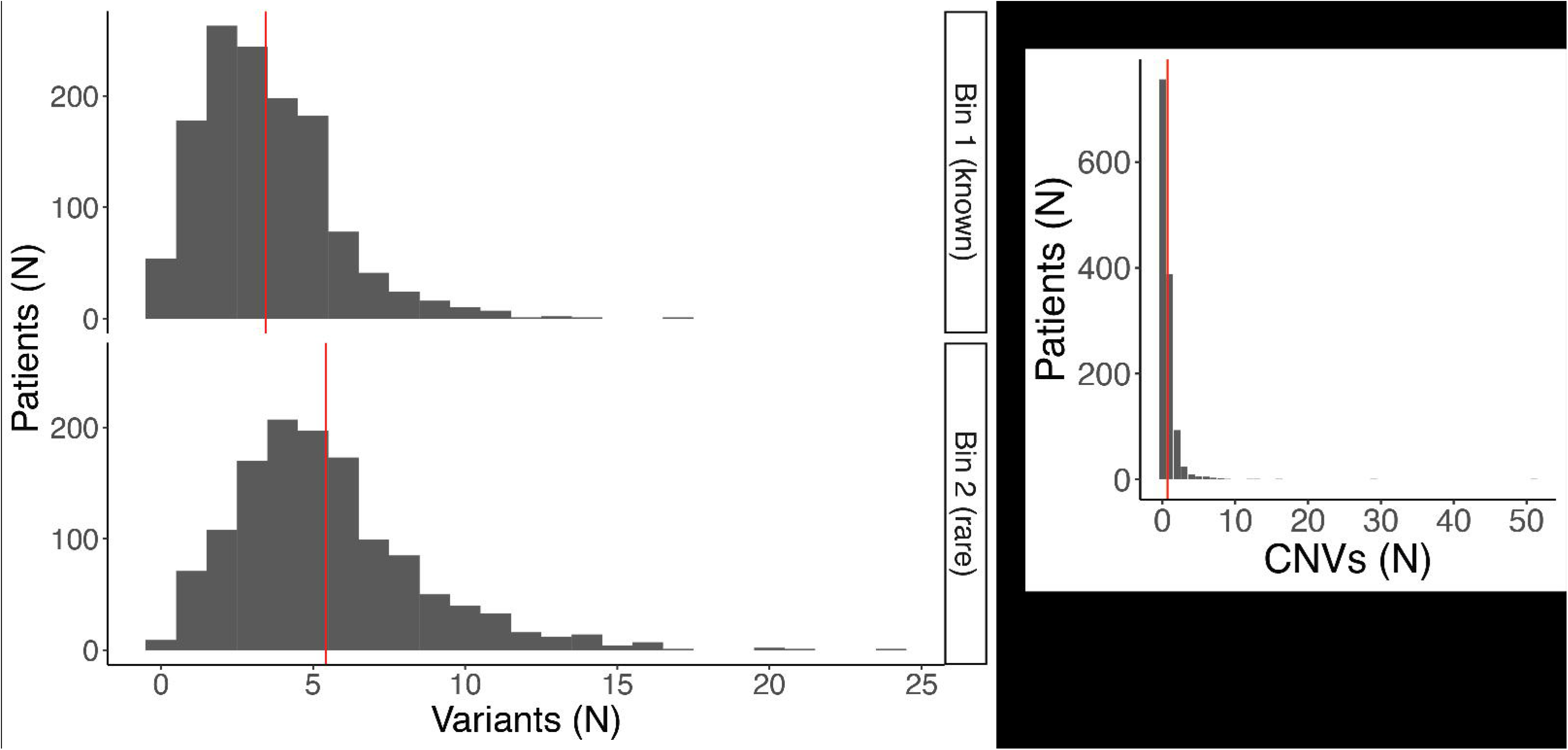
Overview of filtered variants for all clinical exomes. **A** Number of variants in bin 1 (known variants) and bin 2 (rare, non-synonymous coding variants). **B** Amount of copy number variations. The median number of variants per patient is indicated with a red vertical line.

**Figure S4:**
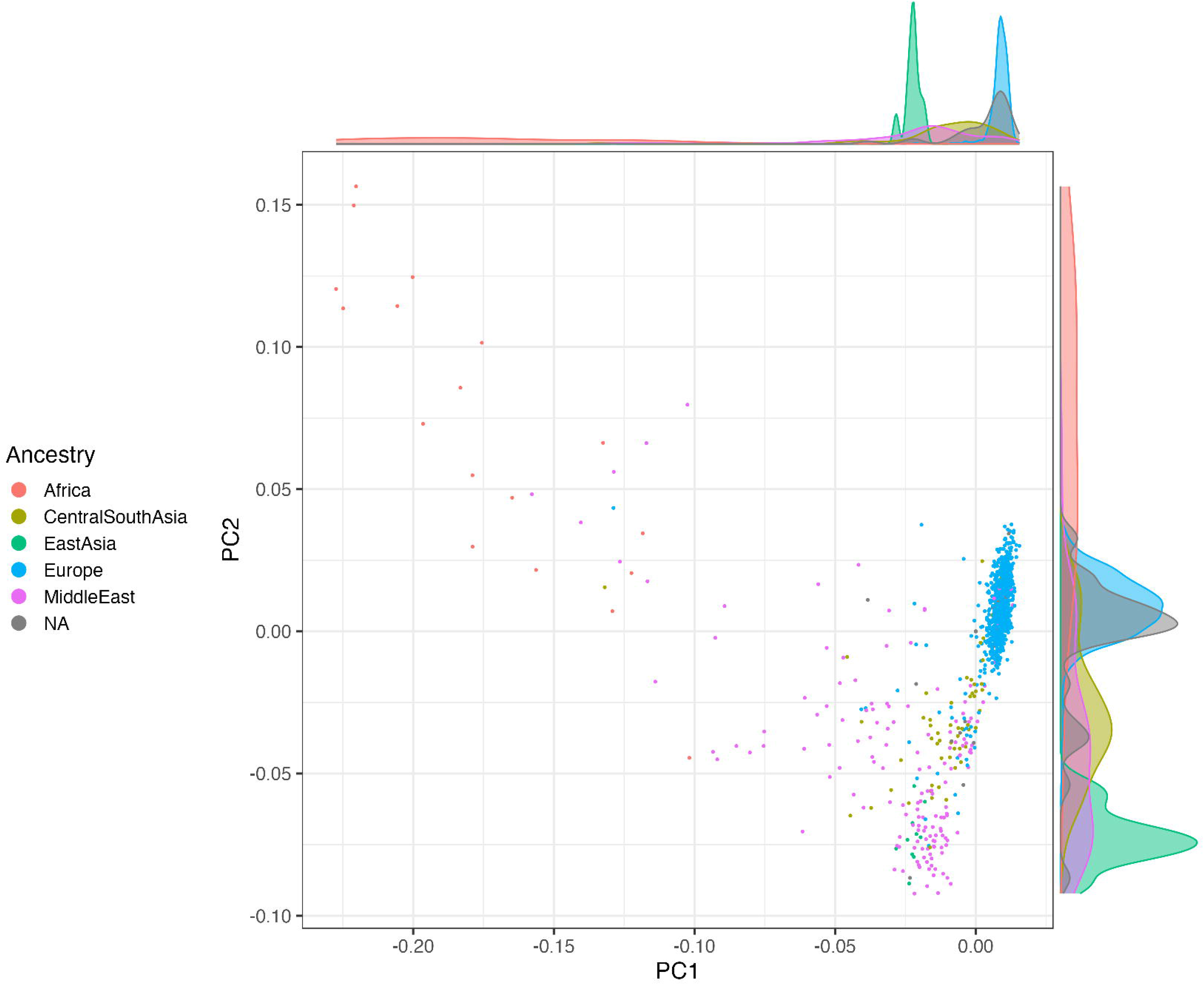
Genetic ancestry analysis of our study population. Principal component analysis was performed and could predict genetic ancestry in 1266 (97.6%) patients in our cohort.

**Figure S5:**
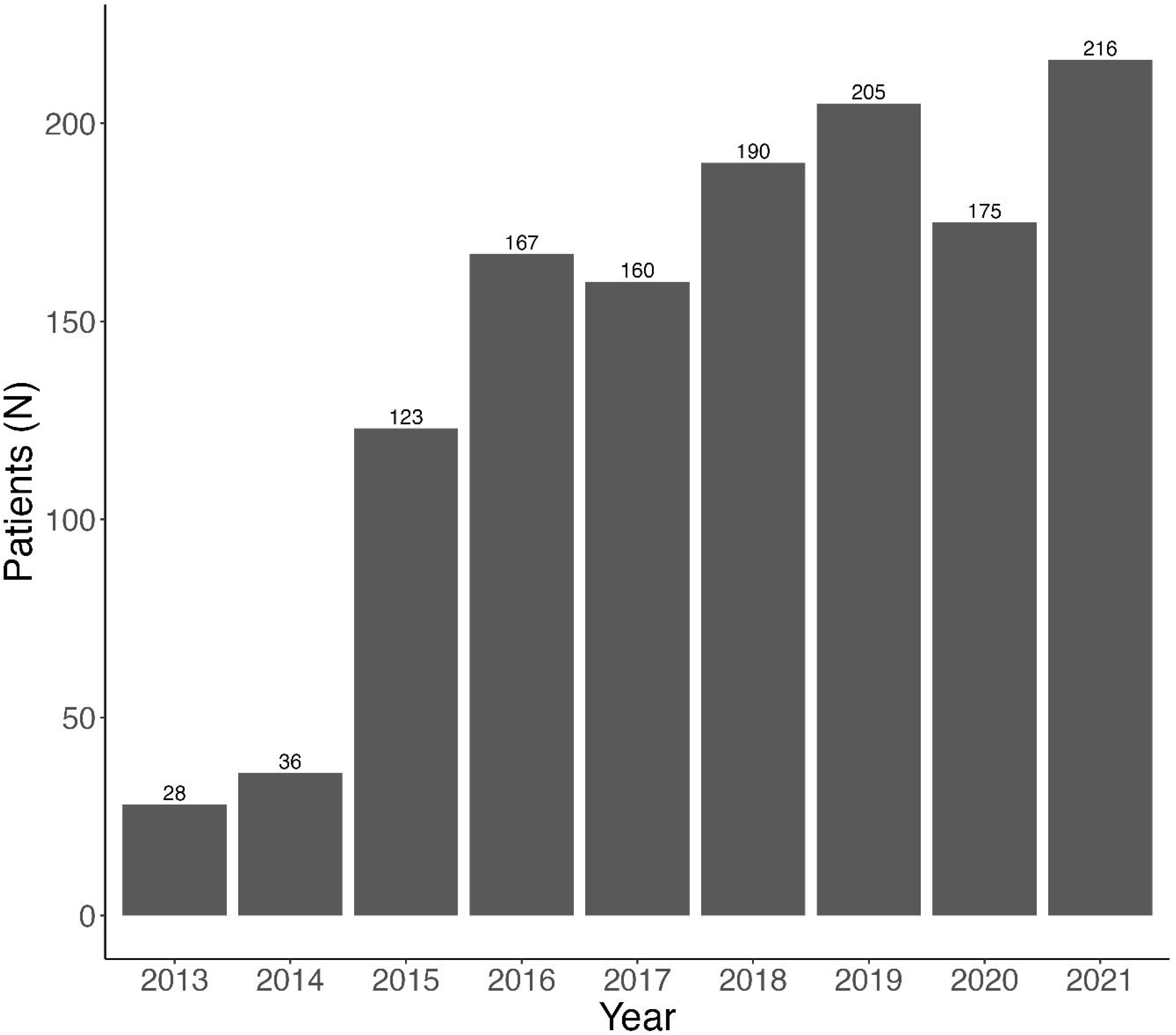
Number of patients included per year. The number of patients in whom the standard analysis was performed per year.

**Figure S6:**
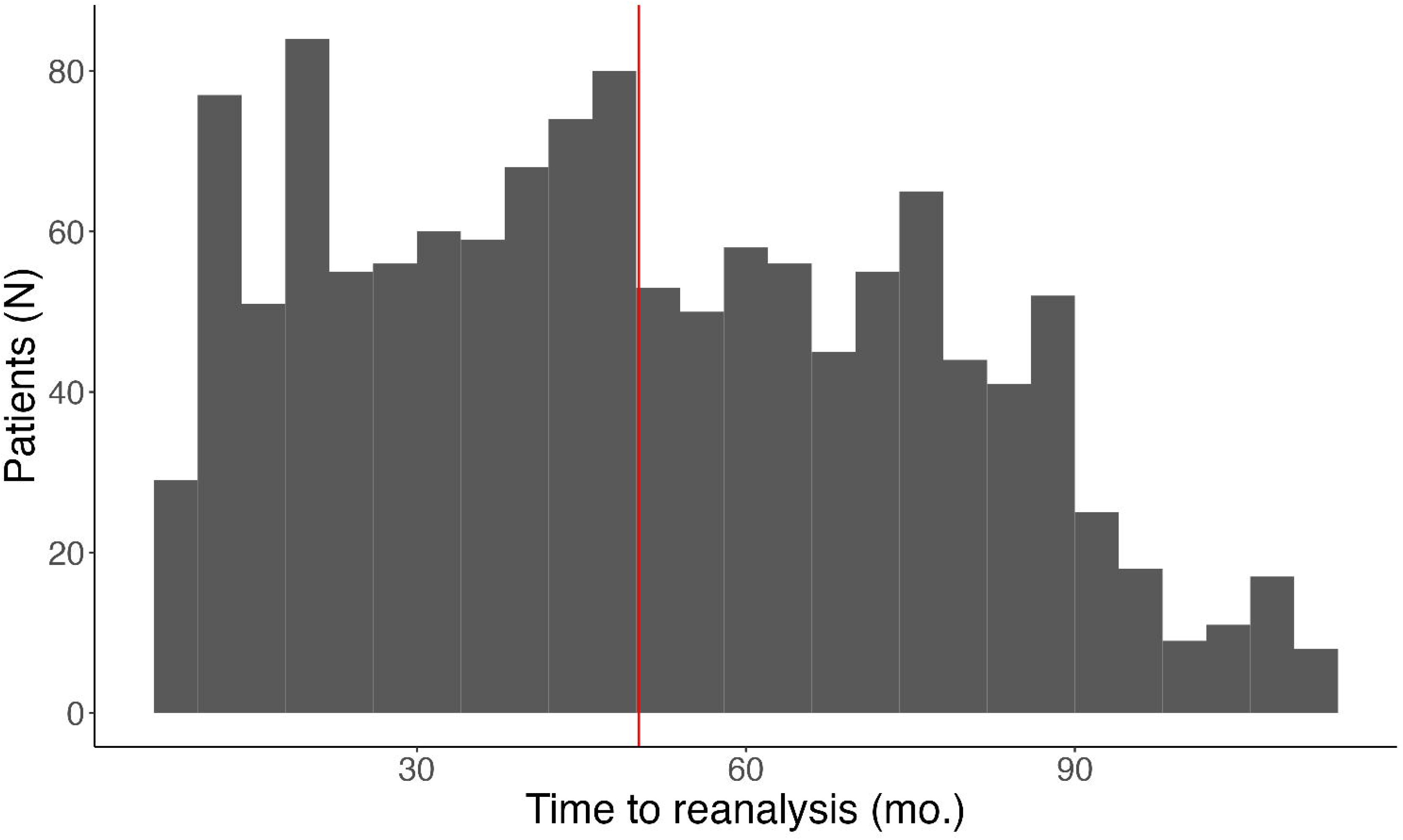
Distribution of the reanalysis interval. Time elapsed between the standard analysis and reanalysis of the inborn errors of immunity patients in our cohort displayed in bins of 4 months. The median number of months is indicated with a red vertical line.

**Figure S7:**
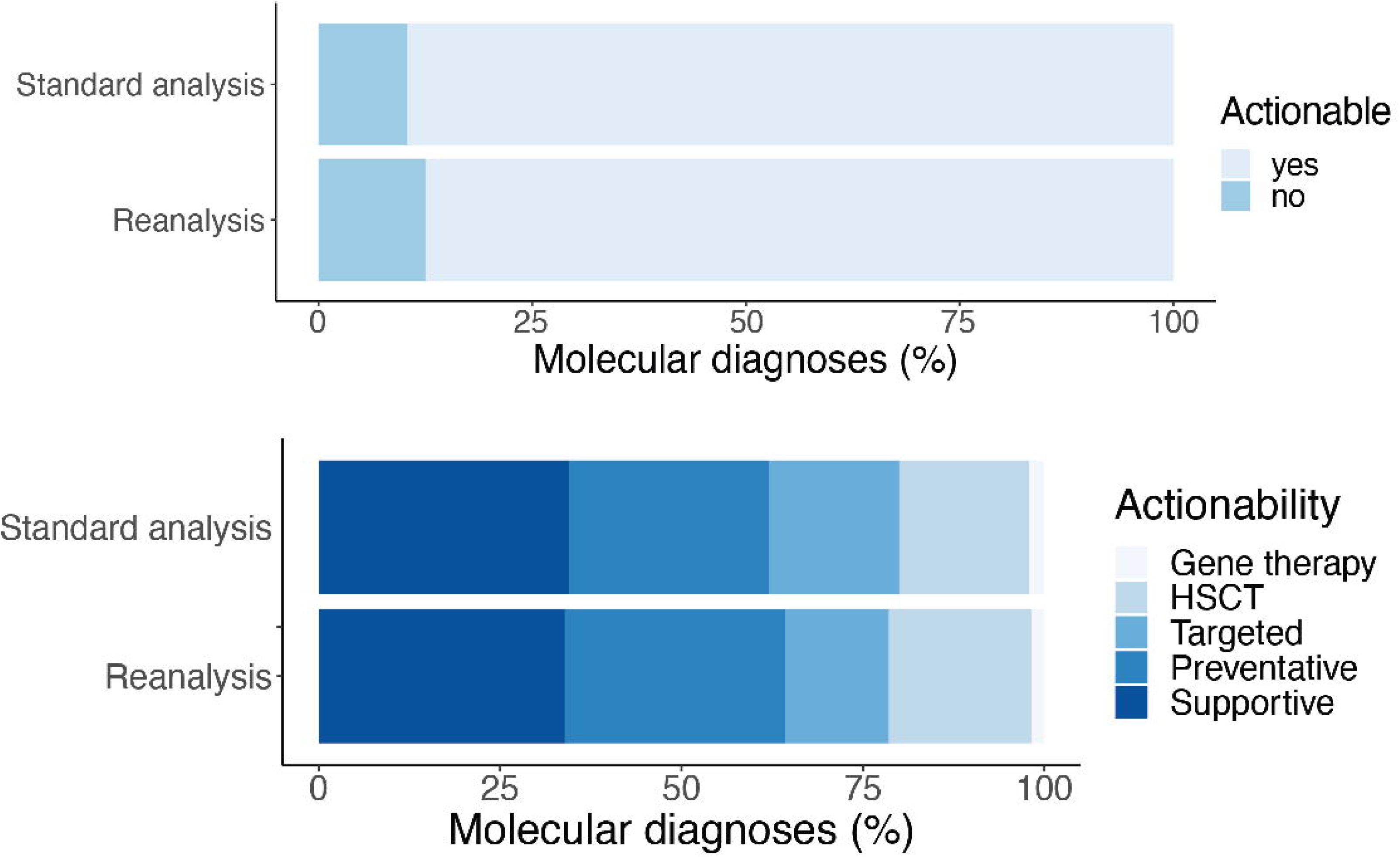
Actionability of genetic diagnoses. Visualization of the data presented in Table S14, which lists options for. actionability associated with each genetic diagnosis identified after standard analysis or reanalysis.

## References

1. Bousfiha A, Moundir A, Tangye SG, Picard C, Jeddane L. The 2022 Update of IUIS Phenotypical Classification for Human Inborn Errors of Immunity. J Clin Immunol. 2022;42:520.

2. Notarangelo LD, Bacchetta R, Casanova JL, Su HC. Human inborn errors of immunity: An expanding universe. Sci Immunol. 2020;5(49).

3. Seth N, Tuano KS, Chinen J. Inborn errors of immunity: Recent progress. J Allergy Clin Immunol. 2021;1–9.

4. Demirdag Y, Fuleihan R, Orange JS, Yu JE. New primary immunodeficiencies 2021 context and future. Curr Opin Pediatr. 2021;33(6):657–75.

5. Arts P, Simons A, Alzahrani MS, Yilmaz E, Alidrissi E, Van Aerde KJ, et al. Exome sequencing in routine diagnostics: A generic test for 254 patients with primary immunodeficiencies. Genome Med. 2019;11(1):1–15.

6. Yska HAF, Elsink K, Kuijpers TW, Frederix GWJ, van Gijn ME, van Montfrans JM. Diagnostic Yield of Next Generation Sequencing in Genetically Undiagnosed Patients with Primary Immunodeficiencies: a Systematic Review. J Clin Immunol. 2019;39(6):577–91.

7. Vorsteveld EE, Hoischen A, Van Der Made CI. Next-Generation Sequencing in the Field of Primary Immunodeficiencies: Current Yield, Challenges, and Future Perspectives. Clin Rev Allergy Immunol. 2021 Oct;61(2):212–25.

8. Wright CF, McRae JF, Clayton S, Gallone G, Aitken S, FitzGerald TW, et al. Making new genetic diagnoses with old data: iterative reanalysis and reporting from genome-wide data in 1,133 families with developmental disorders. Genet Med. 2018;20(10):1216–23.

9. Ewans LJ, Schofield D, Shrestha R, Zhu Y, Gayevskiy V, Ying K, et al. Whole-exome sequencing reanalysis at 12 months boosts diagnosis and is cost-effective when applied early in Mendelian disorders. Genet Med. 2018;20(12):1564–74.

10. Salmon LB, Orenstein N, Markus-Bustani K, Ruhrman-Shahar N, Kilim Y, Magal N, et al. Improved diagnostics by exome sequencing following raw data reevaluation by clinical geneticists involved in the medical care of the individuals tested. Genet Med. 2019;21(6):1443–51.

11. Liu P, Lupski JR, Yang Y. Reanalysis of Clinical Exome Sequencing Data. N Engl J Med. 2019;380(25):2478–80.

12. Tan NB, Stapleton R, Stark Z, Delatycki MB, Yeung A, Hunter MF, et al. Evaluating systematic reanalysis of clinical genomic data in rare disease from single center experience and literature review. Mol Genet Genomic Med. 2020;8(11):1–19.

13. Wenger AM, Guturu H, Bernstein JA, Bejerano G. Systematic reanalysis of clinical exome data yields additional diagnoses: Implications for providers. Genet Med. 2017;19(2):209–14.

14. Schobers G, Schieving JH, Yntema HG, Pennings M, Pfundt R, Derks R, et al. Reanalysis of exome negative patients with rare disease: a pragmatic workflow for diagnostic applications. Genome Med. 2022;14(1):1–10.

15. Mørup SB, Nazaryan-Petersen L, Gabrielaite M, Reekie J, Marquart HV, Hartling HJ, et al. Added Value of Reanalysis of Whole Exome- and Whole Genome Sequencing Data From Patients Suspected of Primary Immune Deficiency Using an Extended Gene Panel and Structural Variation Calling. Front Immunol. 2022 Jun 30;13:906328.

16. Boycott KM, Vanstone MR, Bulman DE, MacKenzie AE. Rare-disease genetics in the era of next-generation sequencing: discovery to translation. Nat Rev Genet. 2013 Oct;14(10):681–91.

17. Boycott KM, Hartley T, Biesecker LG, Gibbs RA, Innes AM, Riess O, et al. A Diagnosis for All Rare Genetic Diseases: The Horizon and the Next Frontiers. Cell. 2019;177(1):32–7.

18. Van Der Made CI, Simons A, Schuurs-Hoeijmakers J, Van Den Heuvel G, Mantere T, Kersten S, et al. Presence of Genetic Variants among Young Men with Severe COVID-19. JAMA - J Am Med Assoc. 2020;324(7):663–73.

19. van der Made CI, Potjewijd J, Hoogstins A, Willems HPJ, Kwakernaak AJ, de Sevaux RGL, et al. Adult-onset autoinflammation caused by somatic mutations in UBA1: A Dutch case series of patients with VEXAS. J Allergy Clin Immunol. 2022 Jan;149(1):432-439.e4.

20. Hebert A, Simons A, Schuurs-Hoeijmakers JH, Koenen HJ, Zonneveld-Huijssoon E, Henriet SS, et al. Trio-based whole exome sequencing in patients with suspected sporadic inborn errors of immunity: A retrospective cohort study. eLife. 2022 Oct 17;11:e78469.

21. Li H, Durbin R. Fast and accurate long-read alignment with Burrows–Wheeler transform. Bioinformatics. 2010 Mar 1;26(5):589–95.

22. McKenna A, Hanna M, Banks E, Sivachenko A, Cibulskis K, Kernytsky A, et al. The Genome Analysis Toolkit: A MapReduce framework for analyzing next-generation DNA sequencing data. Genome Res. 2010 Sep;20(9):1297–303.

23. Krumm N, Sudmant PH, Ko A, O’Roak BJ, Malig M, Coe BP, et al. Copy number variation detection and genotyping from exome sequence data. Genome Res. 2012;22(8):1525–32.

24. Pfundt R, del Rosario M, Vissers LELM, Kwint MP, Janssen IM, de Leeuw N, et al. Detection of clinically relevant copy-number variants by exome sequencing in a large cohort of genetic disorders. Genet Med. 2017 Jun;19(6):667–75.

25. Genome Diagnostics RadboudUMC. PRIMARY IMMUNODEFICIENCY GENE PANEL DG 3.3.0 (482 genes). 2022.

26. Elsink K, Huibers MMH, Hollink IHIM, Simons A, Zonneveld-Huijssoon E, Veken LT van der, et al. Implementation of Early Next-Generation Sequencing for Inborn Errors of Immunityl1: A Prospective Observational Cohort Study of Diagnostic Yield and Clinical Implications in Dutch Genome Diagnostic Centers. Front Immunol. 2021;12:1–11.

27. Stenson PD, Mort M, Ball EV, Evans K, Hayden M, Heywood S, et al. The Human Gene Mutation Database: towards a comprehensive repository of inherited mutation data for medical research, genetic diagnosis and next-generation sequencing studies. Hum Genet. 2017;136(6):665–77.

28. Landrum MJ, Lee JM, Benson M, Brown G, Chao C, Chitipiralla S, et al. ClinVar: Public archive of interpretations of clinically relevant variants. Nucleic Acids Res. 2016;44(D1):D862–8.

29. Wallis Y, Payne S, McAnulty C, Bodmer D, Sistermans E, Robertson K, et al. Practice Guidelines for the Evaluation of Pathogenicity and the Reporting of Sequence Variants in Clinical Molecular Genetics. 2013.

30. Sherry ST, Ward MH, Kholodov M, Baker J, Phan L, Smigielski EM, et al. dbSNP: the NCBI database of genetic variation. Nucleic Acids Res. 2001;29(1):308–11.

31. Karczewski KJ, Weisburd B, Thomas B, Solomonson M, Ruderfer DM, Kavanagh D, et al. The ExAC browser: Displaying reference data information from over 60 000 exomes. Nucleic Acids Res. 2017;45(D1):D840–5.

32. Karczewski KJ, Francioli LC, Tiao G, Cummings BB, Alföldi J, Wang Q, et al. The mutational constraint spectrum quantified from variation in 141,456 humans. Nature. 2020;581(January 2019):434–43.

33. Francioli LC, Menelaou A, Pulit SL, Van Dijk F, Palamara PF, Elbers CC, et al. Whole-genome sequence variation, population structure and demographic history of the Dutch population. Nat Genet. 2014;46(8):818–25.

34. Hamosh A, Scott AF, Amberger JS, Bocchini CA, McKusick VA. Online Mendelian Inheritance in Man (OMIM), a knowledgebase of human genes and genetic disorders. Nucleic Acids Res. 2005;33(DATABASE ISS.):514–7.

35. Pollard KS, Hubisz MJ, Rosenbloom KR, Siepel A. Detection of nonneutral substitution rates on mammalian phylogenies. Genome Res. 2010;20(1):110–21.

36. Kircher M, Witten DM, Jain P, O’roak BJ, Cooper GM, Shendure J. A general framework for estimating the relative pathogenicity of human genetic variants. Nat Genet. 2014;46(3):310–5.

37. Rentzsch P, Witten D, Cooper GM, Shendure J, Kircher M. CADD: Predicting the deleteriousness of variants throughout the human genome. Nucleic Acids Res. 2019;47(D1):D886–94.

38. Richards S, Aziz N, Bale S, Bick D, Das S, Gastier-Foster J, et al. Standards and guidelines for the interpretation of sequence variants: A joint consensus recommendation of the American College of Medical Genetics and Genomics and the Association for Molecular Pathology. Genet Med. 2015;17(5):405–24.

39. Patterson N, Price AL, Reich D. Population Structure and Eigenanalysis. PLoS Genet. 2006;2(12):e190.

40. Matise TC, Chen F, Chen W, De La Vega FM, Hansen M, He C, et al. A second-generation combined linkage–physical map of the human genome: Table 1. Genome Res. 2007 Dec;17(12):1783–6.

41. Wang C, Zhan X, Liang L, Abecasis GR, Lin X. Improved Ancestry Estimation for both Genotyping and Sequencing Data using Projection Procrustes Analysis and Genotype Imputation. Am J Hum Genet. 2015 Jun;96(6):926–37.

42. PRIMARY IMMUNODEFICIENCY GENE PANEL DG 3.5.0 (484 genes). 2022.

43. Sharma M, Leung D, Momenilandi M, Jones LCW, Pacillo L, James AE, et al. Human germline heterozygous gain-of-function STAT6 variants cause severe allergic disease. J Exp Med. 2023 May 1;220(5):e20221755.

44. Hebert A, Simons A, Schuurs-Hoeijmakers JHM, Koenen HJPM, Zonneveld-Huijssoon E, Henriet SSV, et al. Trio-based whole exome sequencing in patients with suspected sporadic inborn errors of immunity: a retrospective cohort study. medRxiv. 2022;1–47.

45. Chen K, Coonrod EM, Kumánovics A, Franks ZF, Durtschi JD, Margraf RL, et al. Germline Mutations in NFKB2 Implicate the Noncanonical NF-kB Pathway in the Pathogenesis of Common Variable Immunodeficiency. Am J Hum Genet. 2013;93(5):812–24.

46. Kleefstra T, Brunner HG, Amiel J, Oudakker AR, Nillesen WM, Magee A, et al. Loss-of-Function Mutations in Euchromatin Histone Methyl Transferase 1 (EHMT1) Cause the 9q34 Subtelomeric Deletion Syndrome. Am J Hum Genet. 2006 Aug;79(2):370–7.

47. Hsu AP, Donkó A, Arrington ME, Swamydas M, Fink D, Das A, et al. Dominant activating RAC2 mutation with lymphopenia, immunodeficiency, and cytoskeletal defects. Blood. 2019 May 2;133(18):1977–88.

48. Smits BM, Lelieveld PHC, Ververs FA, Turkenburg M, de Koning C, van Dijk M, et al. A dominant activating RAC2 variant associated with immunodeficiency and pulmonary disease. Clin Immunol. 2020;212(August 2019):2019–21.

49. van Schouwenburg PA, Davenport EE, Kienzler AK, Marwah I, Wright B, Lucas M, et al. Application of whole genome and RNA sequencing to investigate the genomic landscape of common variable immunodeficiency disorders. Clin Immunol. 2015;160(2):301–14.

50. Nielsen C, Jakobsen MA, Larsen MJ, Müller AC, Hansen S, Lillevang ST, et al. Immunodeficiency Associated with a Nonsense Mutation of IKBKB. J Clin Immunol. 2014;34(8):916–21.

51. Pannicke U, Baumann B, Fuchs S, Henneke P, Rensing-Ehl A, Rizzi M, et al. Deficiency of innate and acquired immunity caused by an IKBKB mutation. N Engl J Med. 2013;369(26):2504–14.

52. Cardinez C, Miraghazadeh B, Tanita K, Silva E, Hoshino A, Okada S, et al. Gain-of-function IKB KB mutation causes human combined immune deficiency. J Exp Med. 2018;215(11):2715–24.

53. Lalaoui N, Boyden SE, Oda H, Wood GM, Stone DL, Chau D, et al. Mutations that prevent caspase cleavage of RIPK1 cause autoinflammatory disease. Nature. 2020 Jan 2;577(7788):103–8.

54. Hunter JE, Irving SA, Biesecker LG, Buchanan A, Jensen B, Lee K, et al. A standardized, evidence-based protocol to assess clinical actionability of genetic disorders associated with genomic variation. Genet Med. 2016 Dec;18(12):1258–68.

55. King JR, Notarangelo LD, Hammarström L. An appraisal of the Wilson & Jungner criteria in the context of genomic-based newborn screening for inborn errors of immunity. J Allergy Clin Immunol. 2021 Feb;147(2):428–38.

56. Pan-Hammarström Q, Salzer U, Du L, Björkander J, Cunningham-Rundles C, Nelson DL, et al. Reexamining the role of TACI coding variants in common variable immunodeficiency and selective IgA deficiency. Nat Genet. 2007 Apr;39(4):429–30.

57. Salzer U, Chapel HM, Webster ADB, Pan-Hammarström Q, Schmitt-Graeff A, Schlesier M, et al. Mutations in TNFRSF13B encoding TACI are associated with common variable immunodeficiency in humans. Nat Genet. 2005;37(8):820–8.

58. Zhang L, Radigan L, Salzer U, Behrens TW, Grimbacher B, Diaz G, et al. Transmembrane activator and calcium-modulating cyclophilin ligand interactor mutations in common variable immunodeficiency: Clinical and immunologic outcomes in heterozygotes. J Allergy Clin Immunol. 2007 Nov;120(5):1178–85.

59. Ji J, Shen L, Bootwalla M, Quindipan C, Tatarinova T, Maglinte DT, et al. A semiautomated whole-exome sequencing workflow leads to increased diagnostic yield and identification of novel candidate variants. Mol Case Stud. 2019 Apr;5(2):a003756.

60. Kakkas I, Tsinti G, Kalala F, Farmaki E, Kourakli A, Kapousouzi A, et al. TACI Mutations in Primary Antibody Deficiencies: A Nationwide Study in Greece. Medicina (Mex). 2021 Aug 16;57(8):827.

61. Bisgin A, Sonmezler O, Boga I, Yilmaz M. The impact of rare and low-frequency genetic variants in common variable immunodeficiency (CVID). Sci Rep. 2021 Apr 15;11(1):8308.

62. Kaplanis J, Samocha KE, Wiel L, Zhang Z, Arvai KJ, Eberhardt RY, et al. Evidence for 28 genetic disorders discovered by combining healthcare and research data. Nature. 2020;(October 2019).

63. Similuk MN, Yan J, Ghosh R, Oler AJ, Franco LM, Setzer M, et al. Clinical Exome Sequencing of 1000 Families with Complex Immune Phenotypes: Towards comprehensive genomic evaluations. J Allergy Clin Immunol. 2022;150(4):54.

64. Stranneheim H, Lagerstedt-robinson K, Magnusson M, Kvarnung M, Nilsson D, Lesko N, et al. Integration of whole genome sequencing into a healthcare settingl1: high diagnostic rates across multiple clinical entities in 3219 rare disease patients. Genome Med. 2021;13(40):1–15.

65. Thaventhiran JED, Lango Allen H, Burren OS, Rae W, Greene D, Staples E, et al. Whole-genome sequencing of a sporadic primary immunodeficiency cohort. Nature. 2020;583:90–5.

66. Salzer U, Grimbacher B. TACI deficiency — a complex system out of balance. Curr Opin Immunol. 2021 Aug;71:81–8.

67. Martínez-Pomar N, Detková D, Arostegui JI, Alvarez A, Soler-Palacín P, Vidaller A, et al. Role of TNFRSF13B variants in patients with common variable immunodeficiency. Blood. 2009 Sep 24;114(13):2846–8.

68. Salzer U, Bacchelli C, Buckridge S, Pan-Hammarström Q, Jennings S, Lougaris V, et al. Relevance of biallelic versus monoallelic TNFRSF13B mutations in distinguishing disease-causing from risk-increasing TNFRSF13B variants in antibody deficiency syndromes. Blood. 2009;113(9):1967–76.

69. Ramirez N, Posadas-Cantera S, Langer N, de Oteyza ACG, Proietti M, Keller B, et al. Multi-omics analysis of naïve B cells of patients harboring the C104R mutation in TACI. Front Immunol. 2022;13(August).

70. Costain G, Jobling R, Walker S, Reuter MS, Snell M, Bowdin S, et al. Periodic reanalysis of whole-genome sequencing data enhances the diagnostic advantage over standard clinical genetic testing. Eur J Hum Genet. 2018 May;26(5):740–4.

71. Nambot S, Thevenon J, Kuentz P, Duffourd Y, Tisserant E, Bruel AL, et al. Clinical whole-exome sequencing for the diagnosis of rare disorders with congenital anomalies and/or intellectual disability: substantial interest of prospective annual reanalysis. Genet Med. 2018 Jun;20(6):645–54.

72. Halfmeyer I, Bartolomaeus T, Popp B, Radtke M, Helms T, Hentschel J, et al. Approach to Cohort-Wide Re-Analysis of Exome Data in 1000 Individuals with Neurodevelopmental Disorders. Genes. 2022 Dec 22;14(1):30.

73. Salfati EL, Spencer EG, Topol SE, Muse ED, Rueda M, Lucas JR, et al. Re-analysis of whole-exome sequencing data uncovers novel diagnostic variants and improves molecular diagnostic yields for sudden death and idiopathic diseases. Genome Med. 2019 Dec;11(1):83.

74. Seo GH, Kim T, Choi IH, Park J young, Lee J, Kim S, et al. Diagnostic yield and clinical utility of whole exome sequencing using an automated variant prioritization system, EVIDENCE. Clin Genet. 2020;98(6):562–70.

75. Matalonga L, Hernández-Ferrer C, Piscia D, Cohen E, Cuesta I, Danis D, et al. Solving patients with rare diseases through programmatic reanalysis of genome-phenome data. Eur J Hum Genet. 2021;29(9):1337–47.

76. Deignan JL, Chung WK, Kearney HM, Monaghan KG, Rehder CW, Chao EC. Points to consider in the reevaluation and reanalysis of genomic test results: a statement of the American College of Medical Genetics and Genomics (ACMG). Genet Med. 2019;21(6):1267–70.

77. Bartolomaeus T, Hentschel J, Jamra RA, Popp B. Re-evaluation and re-analysis of 152 research exomes five years after the initial report reveals clinically relevant changes in 18%. Eur J Hum Genet [Internet]. 2023 Jul 18 [cited 2023 Jul 19]; Available from: https://www.nature.com/articles/s41431-023-01425-6

78. Bowling KM, Thompson ML, Amaral MD, Finnila CR, Hiatt SM, Engel KL, et al. Genomic diagnosis for children with intellectual disability and/or developmental delay. Genome Med. 2017 Dec;9(1):43.

79. Eldomery MK, Coban-Akdemir Z, Harel T, Rosenfeld JA, Gambin T, Stray-Pedersen A, et al. Lessons learned from additional research analyses of unsolved clinical exome cases. Genome Med. 2017 Dec;9(1):26.

80. Kucuk E, van der Sanden BPGH, O’Gorman L, Kwint M, Derks R, Wenger AM, et al. Comprehensive de novo mutation discovery with HiFi long-read sequencing. Genome Med. 2023 May 8;15(1):34.

81. Chaisson MJP, Sanders AD, Zhao X, Malhotra A, Porubsky D, Rausch T, et al. Multi-platform discovery of haplotype-resolved structural variation in human genomes. Nat Commun. 2019 Apr 16;10(1):1784.

82. Lincoln SE, Hambuch T, Zook JM, Bristow SL, Hatchell K, Truty R, et al. One in seven pathogenic variants can be challenging to detect by NGS: an analysis of 450,000 patients with implications for clinical sensitivity and genetic test implementation. Genet Med. 2021;23(9):1673–80.

83. Marwaha S, Knowles JW, Ashley EA. A guide for the diagnosis of rare and undiagnosed disease: beyond the exome. Genome Med. 2022;14(1):1–22.

84. Fusaro M, Coustal C, Barnabei L, Riller Q, Heller M, Ho Nhat D, et al. A large deletion in a non-coding regulatory region leads to NFKB1 haploinsufficiency in two adult siblings. Clin Immunol Orlando Fla. 2024 Apr;261:110165.

85. Hiatt SM, Amaral MD, Bowling KM, Finnila CR, Thompson ML, Gray DE, et al. Systematic reanalysis of genomic data improves quality of variant interpretation. Clin Genet. 2018 Jul;94(1):174–8.

86. Shashi V, Schoch K, Spillmann R, Cope H, Tan QKG, Walley N, et al. A comprehensive iterative approach is highly effective in diagnosing individuals who are exome negative. Genet Med. 2019 Jan;21(1):161–72.

87. Mantere T, Kersten S, Hoischen A. Long-read sequencing emerging in medical genetics. Front Genet. 2019;10(426):1–14.

88. Elsink K, Huibers MMH, Hollink IHIM, van der Veken LT, Ernst RF, Simons A, et al. National external quality assessment for next-generation sequencing-based diagnostics of primary immunodeficiencies. Eur J Hum Genet. 2020;29(1):20–8.

89. Brodszki N, Frazer-Abel A, Grumach AS, Kirschfink M, Litzman J, Perez E, et al. European Society for Immunodeficiencies (ESID) and European Reference Network on Rare Primary Immunodeficiency, Autoinflammatory and Autoimmune Diseases (ERN RITA) Complement Guideline: Deficiencies, Diagnosis, and Management. J Clin Immunol. 2020;40(4):576–91.

90. Zurek B, Ellwanger K, Vissers LELM, Schüle R, Synofzik M, Töpf A, et al. Solve-RD: systematic pan-European data sharing and collaborative analysis to solve rare diseases. Eur J Hum Genet. 2021;

91. Gazzo AM, Daneels D, Cilia E, Bonduelle M, Abramowicz M, Van Dooren S, et al. DIDA: A curated and annotated digenic diseases database. Nucleic Acids Res. 2016;44(D1):D900–7.

92. Papadimitriou S, Gazzo A, Versbraegen N, Nachtegael C, Aerts J, Moreau Y, et al. Predicting disease-causing variant combinations. Proc Natl Acad Sci U S A. 2019;116(24):11878–87.

93. Gruber C, Bogunovic D. Incomplete penetrance in primary immunodeficiency: a skeleton in the closet. Hum Genet. 2020;139(6–7):745–57.

94. Sobreira N, Schiettecatte F, Valle D, Hamosh A. GeneMatcher: A Matching Tool for Connecting Investigators with an Interest in the Same Gene. Hum Mutat. 2015;36(10):928–30.

95. Philippakis AA, Azzariti DR, Beltran S, Brookes AJ, Brownstein CA, Brudno M, et al. The Matchmaker Exchange: A Platform for Rare Disease Gene Discovery. Hum Mutat. 2015;36(10):915–21.

96. Peng XP, Al-Ddafari MS, Caballero-Oteyza A, El Mezouar C, Mrovecova P, Dib SE, et al. Next generation sequencing (NGS)-based approach to diagnosing Algerian patients with suspected inborn errors of immunity (IEIs). Clin Immunol Orlando Fla. 2023 Nov;256:109758.

97. Abolhassani H, Avcin T, Bahceciler N, Balashov D, Bata Z, Bataneant M, et al. Care of patients with inborn errors of immunity in thirty J Project countries between 2004 and 2021. Front Immunol [Internet]. 2022 [cited 2023 Dec 12];13. Available from: https://www.frontiersin.org/articles/10.3389/fimmu.2022.1032358

98. Köhler S, Gargano M, Matentzoglu N, Carmody LC, Lewis-Smith D, Vasilevsky NA, et al. The Human Phenotype Ontology in 2021. Nucleic Acids Res. 2021 Jan 8;49(D1):D1207–17.

