## Supplemental file for "Clinical exome sequencing data from patients with inborn errors of immunity: cohort level meta-analysis and the benefit of systematic reanalysis"

### Reanalysis CNVs

#### Output from CoNIFER

| ID | Chromosome | Start Position | End Position | Length | Type | Value | IEI Gene overlap | Start cytoBand | End cytoBand | Validation | Zygosity |
| --- | --- | --- | --- | --- | --- | --- | --- | --- | --- | --- | --- |
| 155.1 | chr16 | 28996723 | 29096124 | 99402 | del | -2.41 | LAT | p11.2 | p11.2 | Xon array | Homozygous |
| 178.1 | chr16 | 29624260 | 30354548 | 730289 | del | -2.57 | ? | p11.2 | p11.2 | None | Heterozygous (*de novo)* |
| 243.1 | chr10 | 14977403 | 15091811 | 114409 | del | -3.77 | DCLRE1C | p13 | p13 | None | Homozygous |
| 380.1 | chr9 | 1.31E+08 | 1.35E+08 | 3642873 | del | -2.55 | ? | q34.11 | q34.13 | None | Heterozygous |
| 444.1 | chr18 | 74980540 | 77227582 | 2247043 | del | -2.31 | NFATC1 | q23 | q23 | Array | Heterozygous |
| 1145.1 | chr7 | 50070679 | 50436092 | 365414 | DUPLICATION | 2.5 | IKZF1 | p12.2 | p12.2 | None | Heterozygous |

#### Exome coverage at CNV in IGV

**155.1**

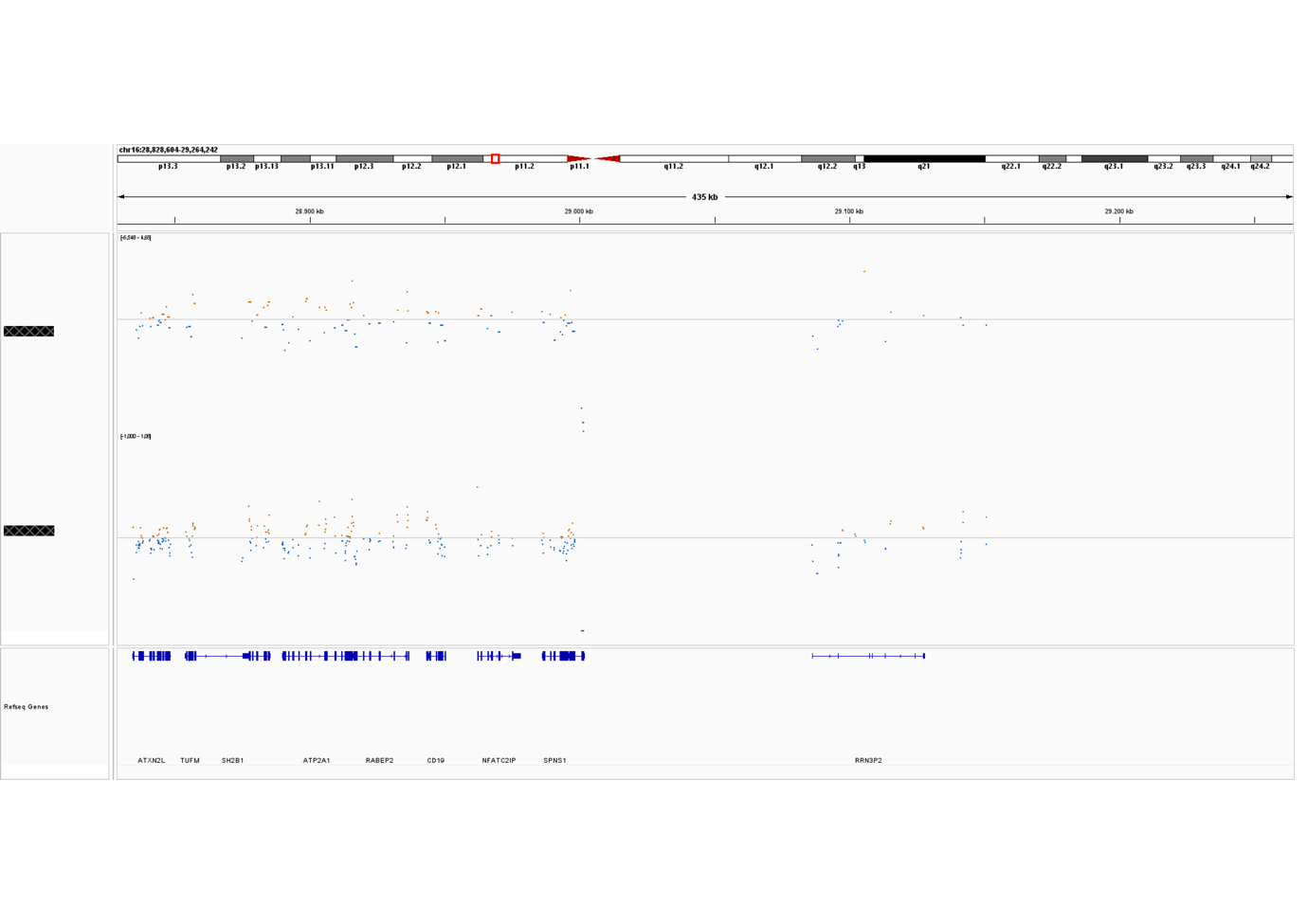

**178.1**

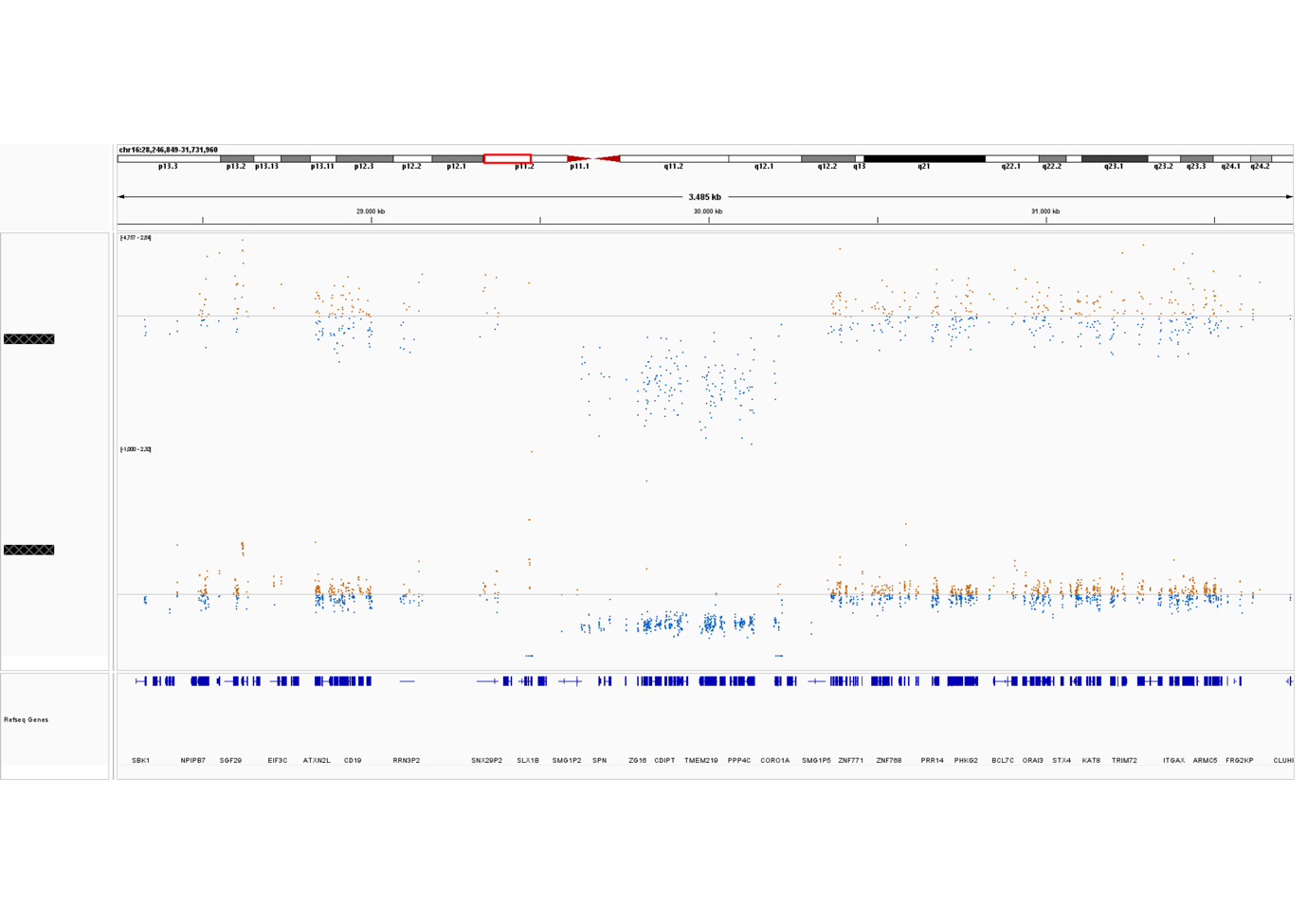

**243.1**

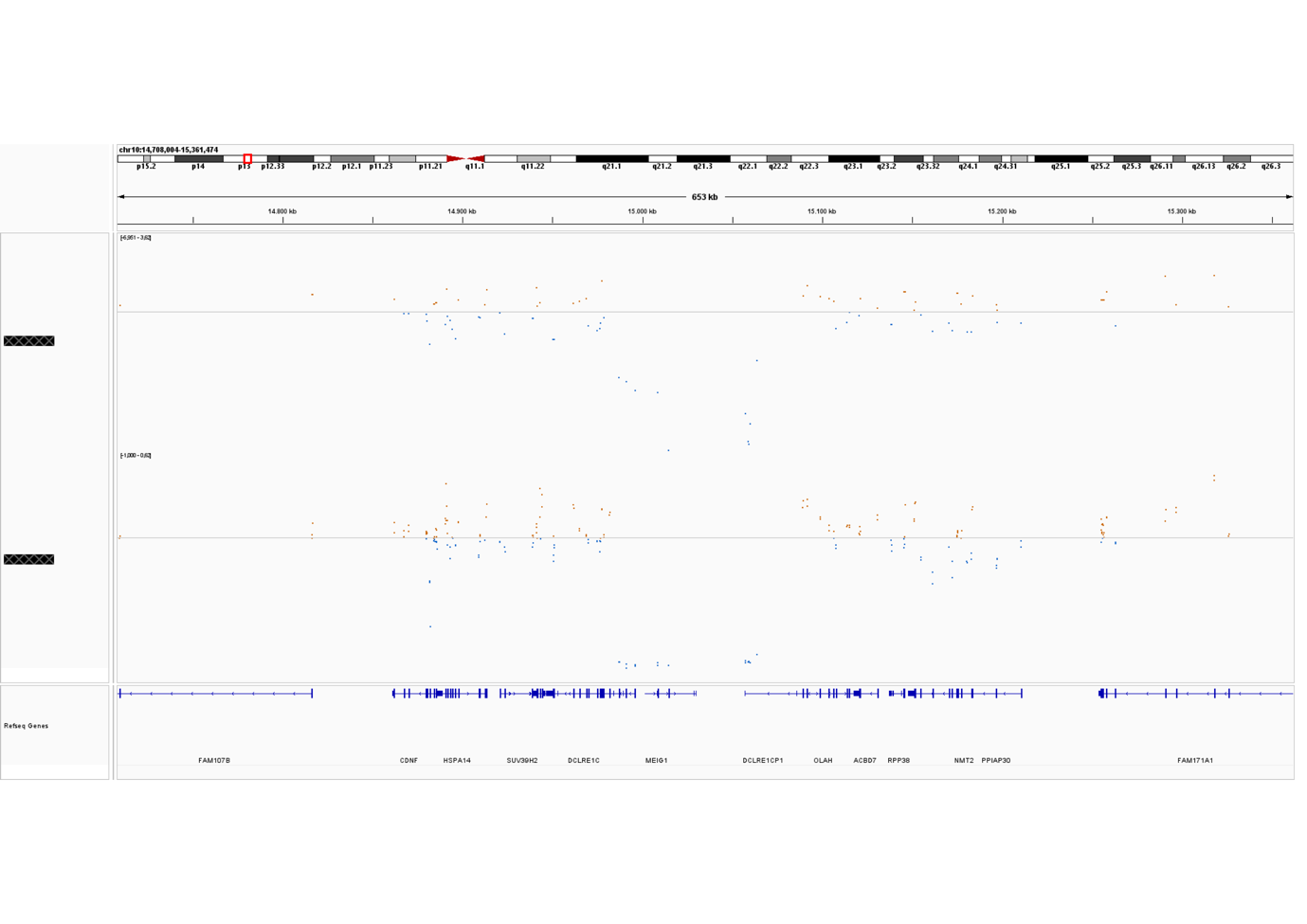

**380.1**

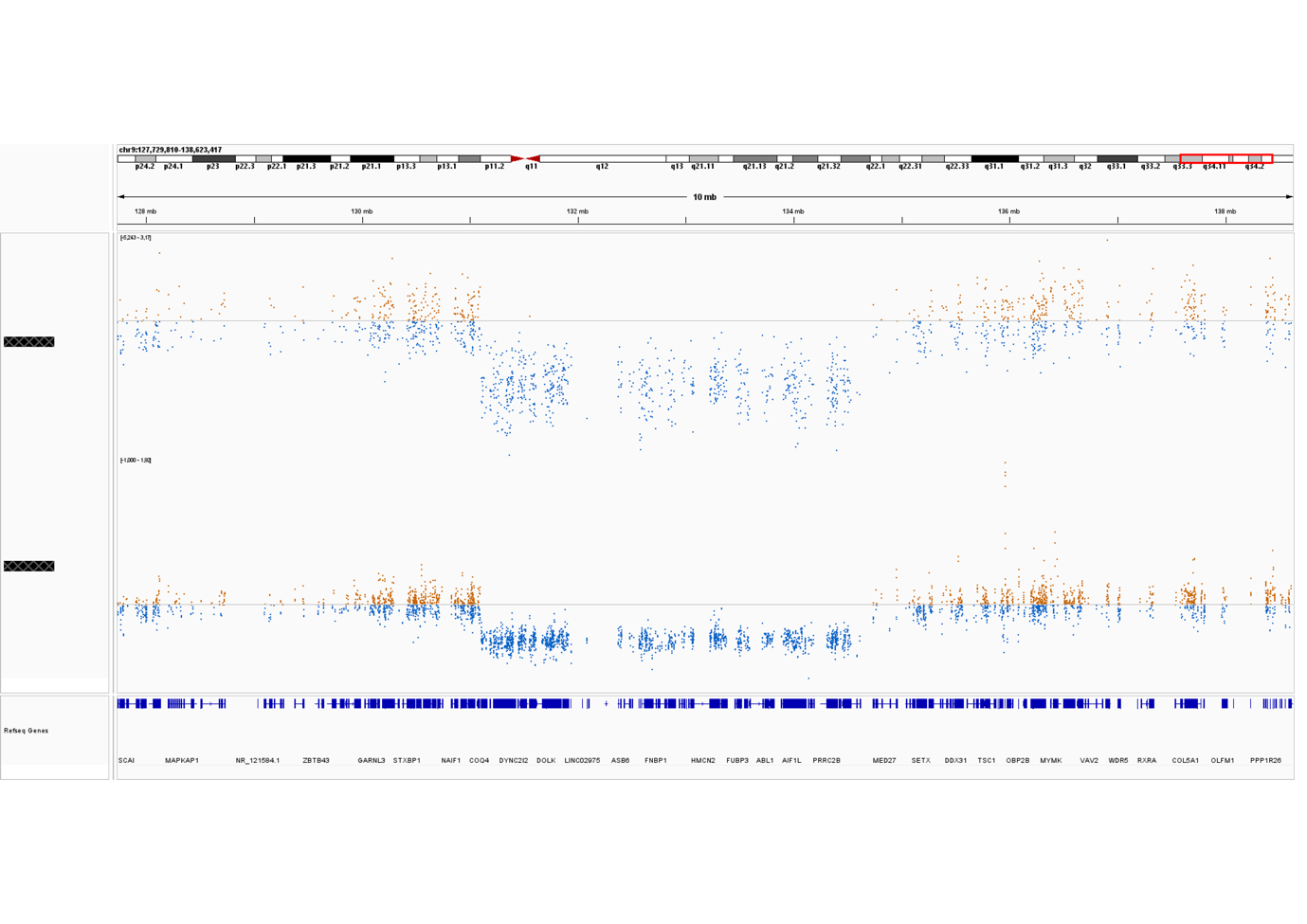

**444.1**

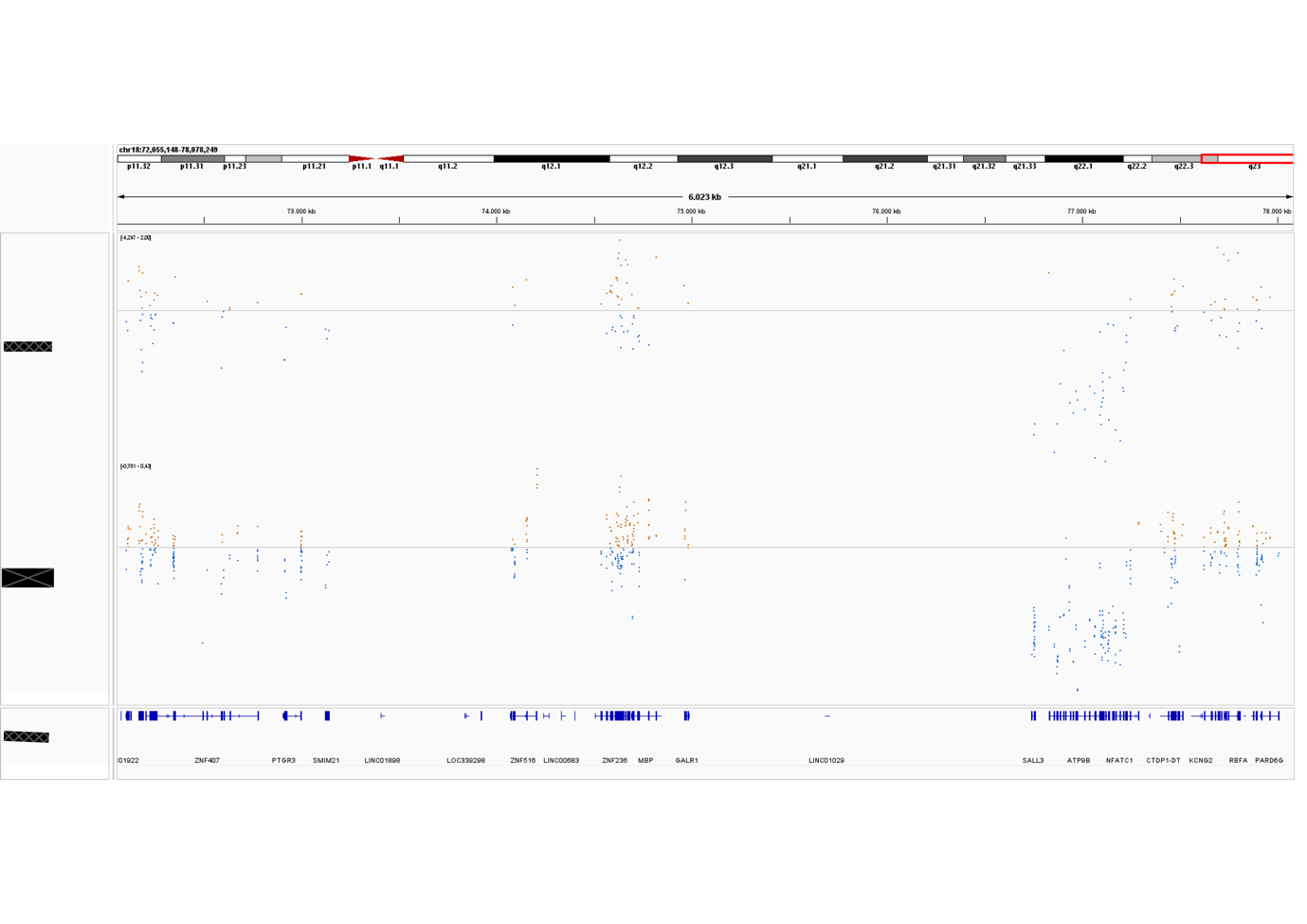

**1145.1**

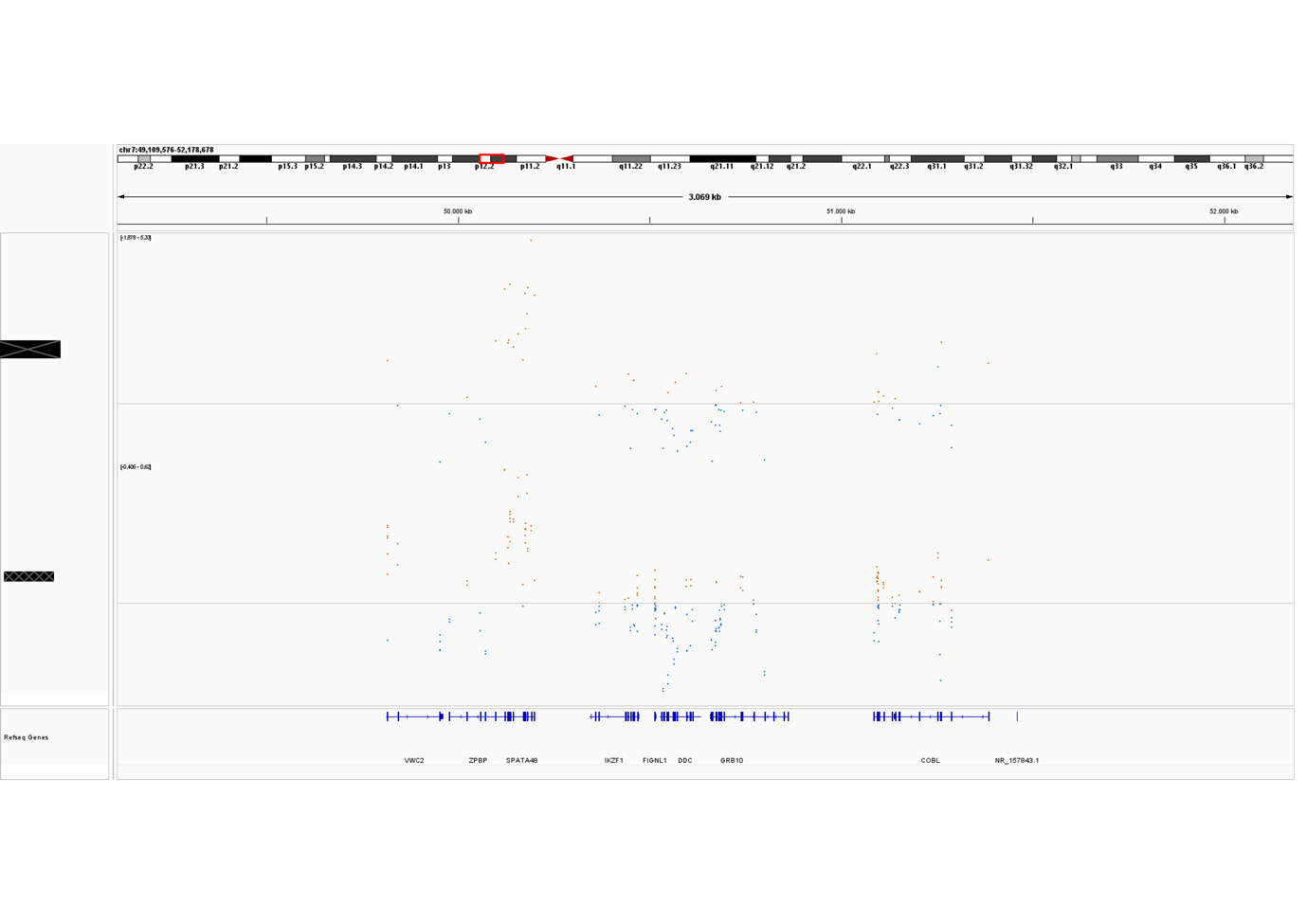
